# Neurometabolic dysfunction in psychosis observed with 7 T MRS

**DOI:** 10.1101/2024.01.03.24300788

**Authors:** Michael-Paul Schallmo, Caroline Demro, Kyle W. Killebrew, Cheryl A. Olman, Scott R. Sponheim, Małgorzata Marjańska

## Abstract

Altered brain chemistry is thought to contribute to impairments in cognitive and perceptual functioning in people with psychotic psychopathology (PwPP). As heritable genetic factors shape the development of psychosis, these alterations in brain chemistry may extend to biological relatives of PwPP. Magnetic resonance spectroscopy (MRS) is a non-invasive method for quantifying the concentration of various neurochemicals in the human brain. A number of MRS studies in different brain regions have been performed in PwPP, and to a lesser extent in relatives, but results have been largely mixed. There are a number of methodological issues that may have influenced previous findings. We show here that when such issues are addressed, MRS reveals a pattern of neurometabolic dysfunction in PwPP. We acquired MRS data at 7 tesla with an ultra-short echo time (TE = 8 ms) sequence in both occipital and prefrontal cortices from 43 healthy controls, 42 first-degree biological relatives, and 64 PwPP. We saw reduced levels of *N*-acetyl-aspartate (NAA) in the occipital lobe in PwPP and their relatives (versus controls), and lower *N*-acetyl-aspartyl-glutamate (NAAG) in prefrontal cortex in PwPP versus controls. Surprisingly, we also saw markedly increased levels of glucose in both occipital and prefrontal cortices in PwPP. Hierarchical clustering analyses showed that higher glucose levels were linked to higher psychiatric symptom levels and impairments in visual task performance. Together, our findings point to a disruption in neural metabolism across multiple brain areas in PwPP that is associated with impaired cognitive and perceptual functioning.

## Introduction

Altered brain chemistry in PwPP is well documented, including altered glutamatergic neurotransmission [2,3], and impaired functioning of the inhibitory neurotransmitter γ-aminobutyric acid (GABA) [4–6], which may reflect a disruption in the balance of excitation and inhibition [7,8]. Beyond neurotransmitter dysfunction, there is also evidence to suggest that psychotic disorders may involve disruptions in brain metabolism [9,10]. However, as many PwPP are prescribed antipsychotic medications that may significantly affect brain chemistry and metabolism, it can be difficult to assess which specific alterations may be causally involved in the etiology of psychosis.

Magnetic resonance spectroscopy (MRS) is a non-invasive method for assessing human brain chemistry. Many MRS studies have been conducted in PwPP, especially in schizophrenia, but published findings have been largely mixed [11–17]. Among metabolites that can be reliably quantified using MRS, those that have shown more consistent differences in PwPP include the neurotransmitters glutamate, GABA, and *N*-acetyl-aspartylglutamate (NAAG), as well as glutamine (involved in glutamate cycling), glutathione (an anti-oxidant), and *N*-acetyl-aspartate (NAA; a marker of neuronal health and metabolic function). A smaller number of MRS studies have been conducted among biological relatives of PwPP [18–22]. Results from these studies generally suggest that brain metabolite levels in relatives fall on a continuum between healthy controls and PwPP.

There are several methodological challenges that must be considered for MRS studies. These include low concentrations in the brain for many metabolites of interest (e.g., GABA, ∼ 1 mM [23]), and a high degree of spectral overlap (e.g., for glutamate and glutamine), necessitating methods with sufficiently high signal-to-noise ratio (SNR) and spectral resolution. There are also issues with quantification methods, such as the need to properly account for signals from macromolecules that may overlap resonances from metabolites of interest [24], as well as adopting modeling techniques (e.g., using a rigid baseline) [25,26] to avoid over-fitting and quantification errors. Finally, there is a concern that transverse relaxation time constants (*T_2_*) for some metabolites (e.g., NAA) may be shorter in PwPP [27–30]. If so, this may confound quantification of metabolite concentrations; at sufficiently long echo times (TEs), shorter *T_2_* would lead to smaller metabolite signals in PwPP, even in the absence of any true difference in concentration. These issues have been inconsistently addressed across prior MRS studies of PwPP.

The goal of the current study was to use state of the art methods to quantify abnormalities in brain metabolite levels in PwPP using 7 tesla MRS. Acquiring our data at 7 T provided us with higher SNR and improved our ability to study specific metabolites of interest (i.e., separately quantifying glutamate and glutamine, reliably measuring GABA), as compared to similar 3 T methods [31–35]. We explicitly accounted for macromolecules in our basis set, fitted our metabolites without soft constraints, and used a rigid baseline during fitting, to avoid quantification errors [24–26,36]. We also used an ultra-short TE (8 ms) to mitigate the potential confound of shorter *T_2_* in PwPP.

Because perceptual abnormalities are a defining characteristic of psychosis, we are particularly interested in how disrupted visual perception is in PwPP. Thus, we chose a mid-occipital volume of interest (VOI) that included areas such as primary visual cortex, as this part of the brain is thought to play a key role in a number of fundamental visual functions including coding for contrast, orientation, color, motion, and spatial frequency [37]. Our secondary interest was in examining neurochemical markers in prefrontal regions that may be associated with higher-level cognitive or perceptual impairments in people with psychosis. Thus, we chose to examine a second VOI within the dorsomedial prefrontal cortex, as this region is thought to play a role in functions such as decision making and cognitive control [38–40]. We chose to take a transdiagnostic approach to studying psychosis, as the reliability and validity of traditional psychiatric diagnostic categories (e.g., schizophrenia vs. bipolar disorder) have been criticized [41,42]. In addition to PwPP and healthy controls, we also chose to examine MRS markers from first-degree biological relatives of PwPP, who on average share 50% of their genes with someone with psychosis. This allowed us to examine the role of genetic liability for psychosis in brain chemistry, in the absence of antipsychotic medication effects.

## Methods

Our experimental methods are described in full in our recent publications [1,43]. Essential methodological information for the current study is provided below.

### Participants

We collected MRS data from three groups: healthy control participants with no family history of psychosis (controls, n = 44), first-degree biological relatives of individuals with psychosis (relatives, n = 44), and people with a history of psychotic psychopathology (PwPP, n = 66). Of these, 5 individuals (1 control, 2 relatives, 2 PwPP) were excluded for eligibility reasons (see below) and are not considered further. Our final sample included 43 controls, 42 relatives, and 64 PwPP. Demographic information is presented in Table 1.

**Table 1.**
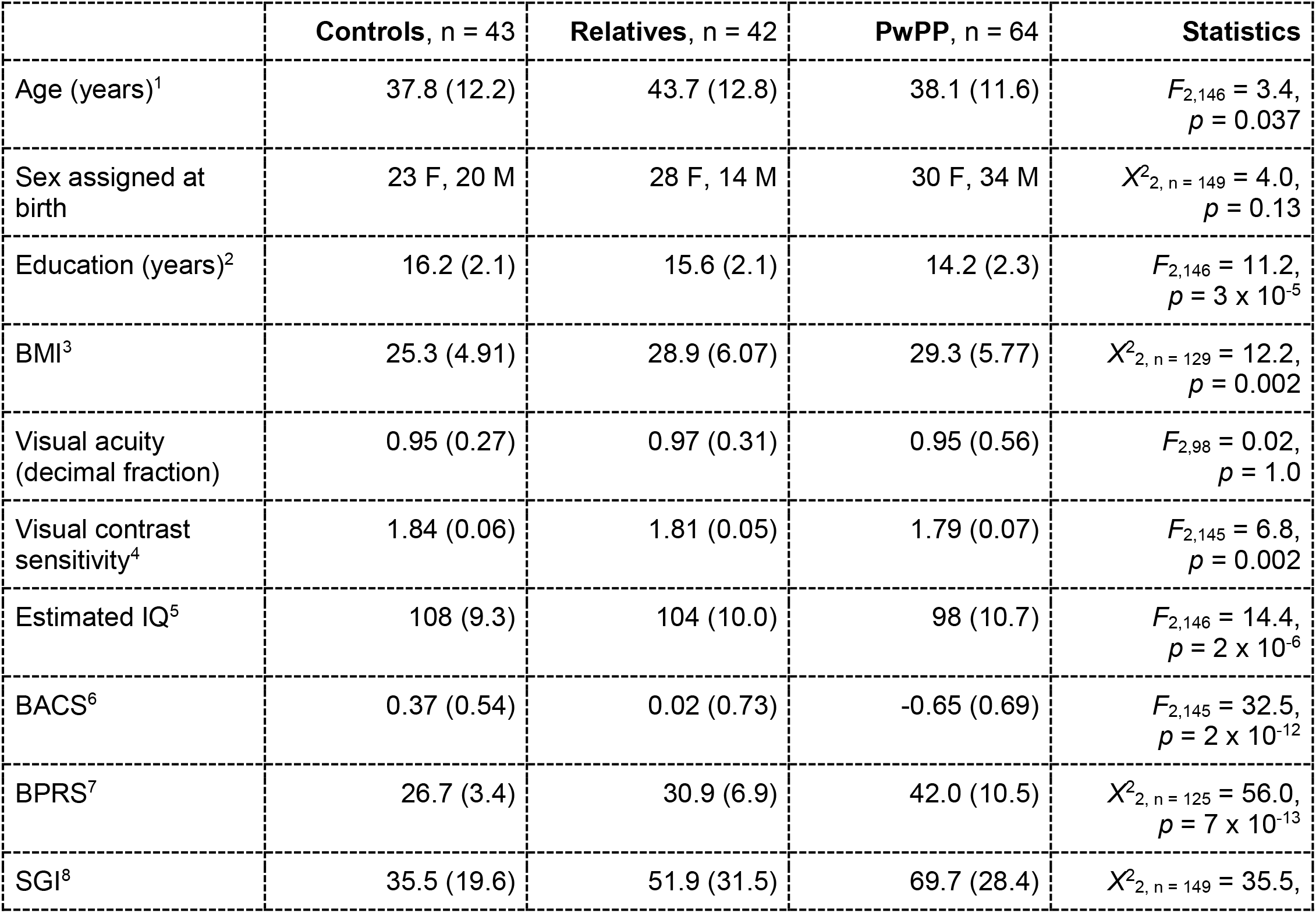
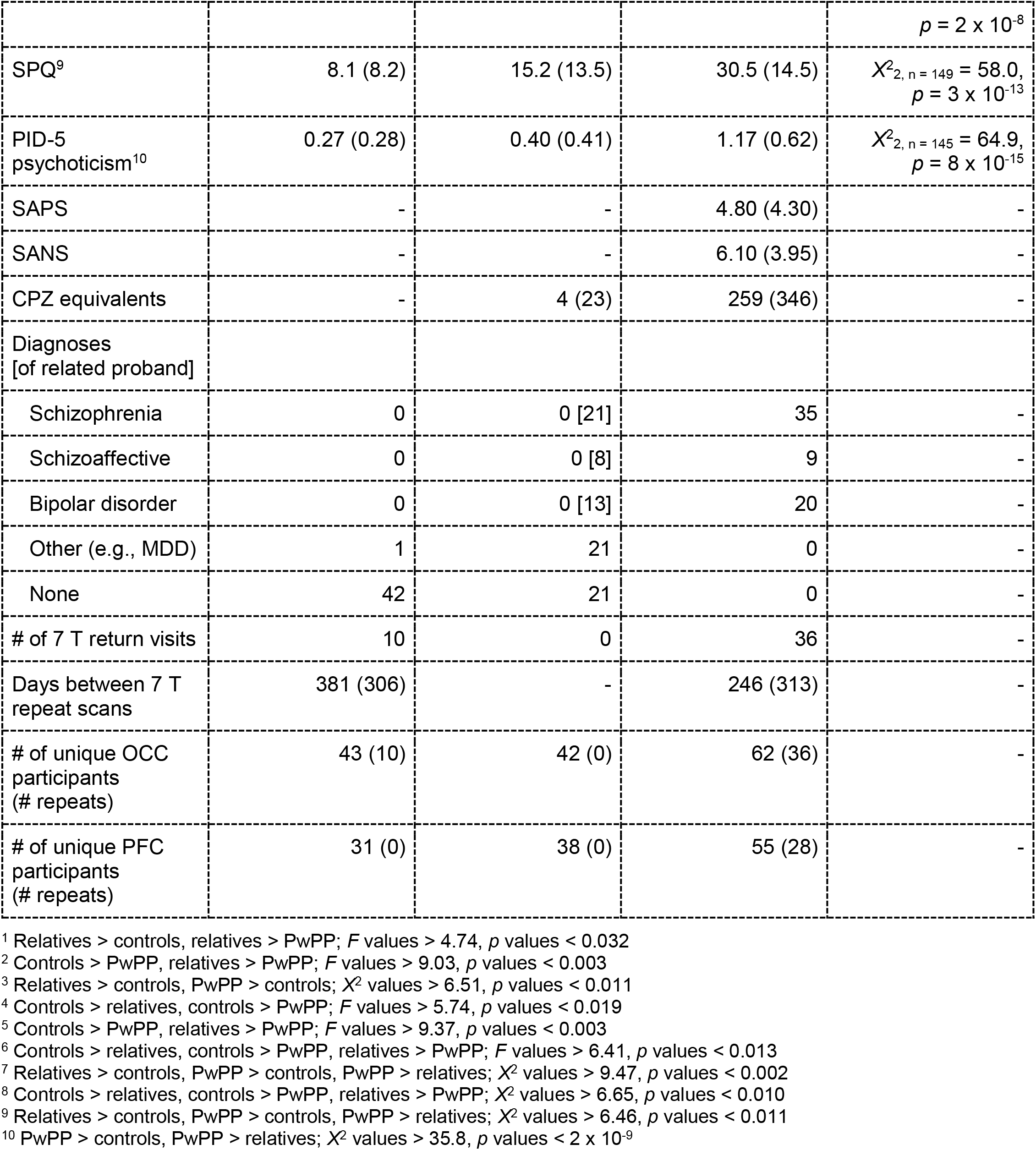
Participant demographics. Data are mean (*SD*) unless otherwise noted. The statistics column shows the test statistic value and *p value* for differences across all three groups. Superscript numbers denote significant *post-hoc* differences between groups at *p* < 0.05, as noted below. For measures where normality or homogeneity of variance were not observed, non-parametric Kruskal-Wallis tests (*Χ*^2^-values) were used instead of analyses of variance (*F*-values). BMI = body mass index. Visual acuity was assessed with a Snellen eye chart [44]; the decimal fraction is reported (e.g., 0.5 indicates 20/40). Visual contrast sensitivity was assessed with the Mars Letter Contrast Sensitivity test [45]. Estimated IQ was assessed using the Wechsler Adult Intelligence Scale [WAIS-IV; 46]. BACS = Brief Assessment of Cognition in Schizophrenia, Z-score [47]. BPRS = Brief Psychiatric Rating Scale Total Score [48]. SGI = Sensory Gating Inventory [49]. SPQ = Schizotypal Personality Questionnaire [50]. PID-5 psychoticism = psychoticism factor from the Personality Inventory for DSM-5 [51]. SANS = Scale for the Assessment of Negative Symptoms Total Global Score [52]. SAPS = Scale for the Assessment of Positive Symptoms Total Global Score [53]. CPZ = chlorpromazine dose equivalents, using established methods [54,55]. Diagnoses were based on the Structured Clinical Interview for DSM-IV-TR disorders [SCID; 56]. For relatives, under Diagnoses, the number of related individuals with a particular psychotic disorder diagnosis is listed in square brackets. MDD = Major Depressive Disorder. OCC = Occipital cortex. PFC = Prefrontal cortex.

| | Controls, $n = 43$ | Relatives, $n = 42$ | PwPP, $n = 64$ | Statistics |
| --- | --- | --- | --- | --- |
| Age (years)<sup>1</sup> | 37.8 (12.2) | 43.7 (12.8) | 38.1 (11.6) | $F_{2,146} = 3.4$,<br>$p = 0.037$ |
| Sex assigned at birth | 23 F, 20 M | 28 F, 14 M | 30 F, 34 M | $X^2_{2, n = 149} = 4.0$,<br>$p = 0.13$ |
| Education (years)<sup>2</sup> | 16.2 (2.1) | 15.6 (2.1) | 14.2 (2.3) | $F_{2,146} = 11.2$,<br>$p = 3 \times 10^{-5}$ |
| BMI<sup>3</sup> | 25.3 (4.91) | 28.9 (6.07) | 29.3 (5.77) | $X^2_{2, n = 129} = 12.2$,<br>$p = 0.002$ |
| Visual acuity (decimal fraction) | 0.95 (0.27) | 0.97 (0.31) | 0.95 (0.56) | $F_{2,98} = 0.02$,<br>$p = 1.0$ |
| Visual contrast sensitivity<sup>4</sup> | 1.84 (0.06) | 1.81 (0.05) | 1.79 (0.07) | $F_{2,145} = 6.8$,<br>$p = 0.002$ |
| Estimated IQ<sup>5</sup> | 108 (9.3) | 104 (10.0) | 98 (10.7) | $F_{2,146} = 14.4$,<br>$p = 2 \times 10^{-6}$ |
| BACS<sup>6</sup> | 0.37 (0.54) | 0.02 (0.73) | -0.65 (0.69) | $F_{2,145} = 32.5$,<br>$p = 2 \times 10^{-12}$ |
| BPRS<sup>7</sup> | 26.7 (3.4) | 30.9 (6.9) | 42.0 (10.5) | $X^2_{2, n = 125} = 56.0$,<br>$p = 7 \times 10^{-13}$ |
| SGI<sup>8</sup> | 35.5 (19.6) | 51.9 (31.5) | 69.7 (28.4) | $X^2_{2, n = 149} = 35.5$, |
| | | | | $p = 2 \times 10^{-8}$ |
| SPQ<sup>9</sup> | 8.1 (8.2) | 15.2 (13.5) | 30.5 (14.5) | $X^2_{2, n=149} = 58.0$,<br>$p = 3 \times 10^{-13}$ |
| PID-5<br>psychoticism<sup>10</sup> | 0.27 (0.28) | 0.40 (0.41) | 1.17 (0.62) | $X^2_{2, n=145} = 64.9$,<br>$p = 8 \times 10^{-15}$ |
| SAPS | - | - | 4.80 (4.30) | - |
| SANS | - | - | 6.10 (3.95) | - |
| CPZ equivalents | - | 4 (23) | 259 (346) | - |
| Diagnoses<br>[of related proband] |  |  |  |  |
| Schizophrenia | 0 | 0 [21] | 35 | - |
| Schizoaffective | 0 | 0 [8] | 9 | - |
| Bipolar disorder | 0 | 0 [13] | 20 | - |
| Other (e.g., MDD) | 1 | 21 | 0 | - |
| None | 42 | 21 | 0 | - |
| # of 7 T return visits | 10 | 0 | 36 | - |
| Days between 7 T<br>repeat scans | 381 (306) | - | 246 (313) | - |
| # of unique OCC<br>participants<br>(# repeats) | 43 (10) | 42 (0) | 62 (36) | - |
| # of unique PFC<br>participants<br>(# repeats) | 31 (0) | 38 (0) | 55 (28) | - |

<sup>1</sup> Relatives > controls, relatives > PwPP; $F$ values > 4.74, $p$ values < 0.032
<sup>2</sup> Controls > PwPP, relatives > PwPP; $F$ values > 9.03, $p$ values < 0.003
<sup>3</sup> Relatives > controls, PwPP > controls; $X^2$ values > 6.51, $p$ values < 0.011
<sup>4</sup> Controls > relatives, controls > PwPP; $F$ values > 5.74, $p$ values < 0.019
<sup>5</sup> Controls > PwPP, relatives > PwPP; $F$ values > 9.37, $p$ values < 0.003
<sup>6</sup> Controls > relatives, controls > PwPP, relatives > PwPP; $F$ values > 6.41, $p$ values < 0.013
<sup>7</sup> Relatives > controls, PwPP > controls, PwPP > relatives; $X^2$ values > 9.47, $p$ values < 0.002
<sup>8</sup> Controls > relatives, controls > PwPP, relatives > PwPP; $X^2$ values > 6.65, $p$ values < 0.010
<sup>9</sup> Relatives > controls, PwPP > controls, PwPP > relatives; $X^2$ values > 6.46, $p$ values < 0.011
<sup>10</sup> PwPP > controls, PwPP > relatives; $X^2$ values > 35.8, $p$ values < $2 \times 10^{-9}$

A subset of individuals (10 controls, 36 PwPP) returned for an identical repeat scanning session several months after their initial MRS study visit. This permitted us to examine longitudinal variability in MRS metabolite measures. Data regarding repeat scans are included in Table 1. Because we obtained repeat scan data from a subset of participants, we refer to data acquired from one participant in a single scanning session as a ‘data set,’ and there are more data sets than unique participants in this study.

Our inclusion and exclusion criteria have been described previously [1,43], and are provided in the Supplemental information. Relatives included people with and without current psychiatric diagnoses, but none with psychosis. All participants provided written informed consent prior to participation, and were compensated $20 per hour. Our procedures complied with the Declaration of Helsinki and were approved by the Institutional Review Board at the University of Minnesota.

### Apparatus

We acquired 7 tesla MRS data using a Siemens MAGNETOM scanner equipped with an 8-kW RF power amplifier and body gradients with 70 mT/m maximum amplitude and 200 T/m/s maximum slew rate. We used a custom-built radio frequency (RF) head coil with two surface ^1^H quadrature transceivers, one for the occipital cortex and one for the prefrontal cortex. Additional details are provided in the Supplemental information.

### Experimental protocol

#### MR spectroscopy

The following information conforms to established MRS reporting standards [1,57]. We used a simulated echo acquisition mode (STEAM) sequence with repetition time (TR) = 5 s, echo time (TE) = 8 ms (see Supplemental information for additional details). For each volume-of-interest (VOI), we acquired 96 shots with a scan duration of 8 min. We also acquired a water reference scan for each VOI (1 average, transmitter frequency = 4.7 ppm, no water suppression) for quantifying absolute metabolite concentrations relative to water, and to facilitate eddy current compensation. We used FAST(EST)MAP [58,59] to perform shimming within each VOI in order to obtain a water linewidth ≤ 15 Hz.

The occipital VOI size was 30 mm (left-right) x 18 mm (anterior-posterior) x 18 mm (inferior-superior). We positioned the VOI within the medial occipital lobe, superior to the cerebellar tentorium, anterior to the occipital pole, and posterior to the occipitoparietal junction (Figure 1). This VOI encompasses regions including primary visual cortex that are known to play a role in fundamental aspects of visual perception. We chose to align and position our VOI squarely in the center of the occipital lobe, in order to cover the region of maximum transmit power efficiency of our RF coil, and to promote inter-operator reliability for positioning. Repeat scans were positioned automatically using the VOI placement information from the first scanning session, via Siemens AutoAlign.

**Figure 1.**
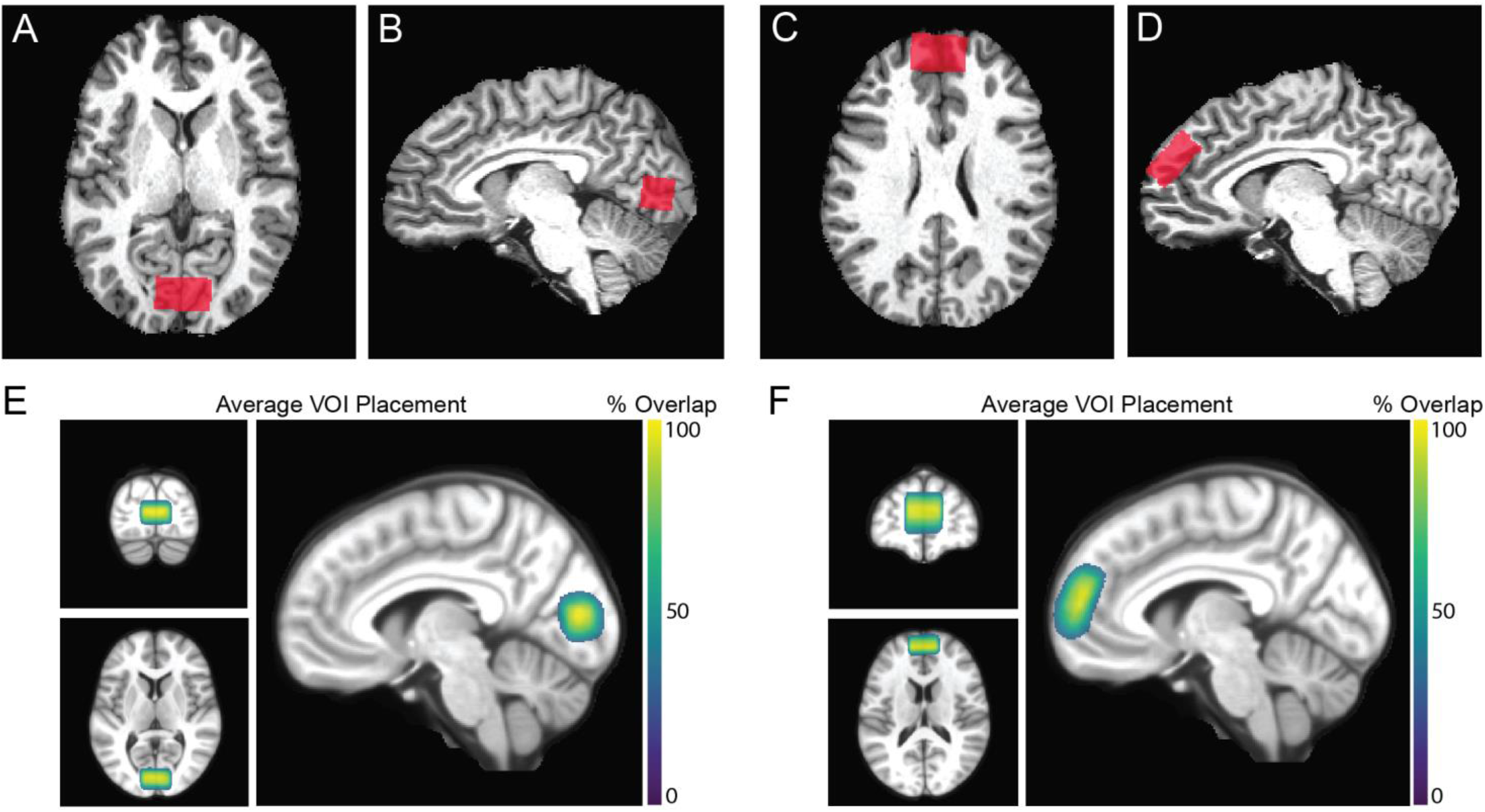
Occipital and prefrontal volumes of interest (VOIs). A) Axial and B) sagittal images from an individual participant showing occipital VOI positioning. Panels C) and D) show the same, but for the prefrontal VOI. E) Average occipital VOI placement across 193 data sets, including repeat scans. Color bar shows % overlap across data sets. Voxels with ≥ 30% overlap are shown. VOIs were transformed from individual to MNI space. F) Same, but for the prefrontal VOI, from 147 data sets. Figures adapted from [1].

The PFC VOI size was 30 mm (left-right) x 30 mm (anterior-posterior) x 15 mm (inferior-superior). This VOI was obliquely oriented within the dorsomedial prefrontal cortex, superior to the anterior cingulate, along a line extending 135° from the anterior edge of the genu of the corpus callosum (Figure 1). We collected 128 or 96 shots for the PFC (see Supplemental information). Total MRS scanning time for both VOIs combined was approximately 80 min.

#### Visual tasks

Our visual paradigms are described in the Supplemental Information and our recent publication [1].

#### Clinical measures

Our clinical research methods are described in the Supplemental Information and our recent publications [1,43].

### Data processing & quality assessment

MRS data processing included eddy current compensation for the metabolite spectra and our water reference scan. We then performed frequency and phase correction, as previously described [1]. Additional details are provided in the Supplemental information.

We quantified metabolite concentrations (Figure 2) using LCModel version 6.3-1N [60]. We used a basis set obtained from a previous study [61], which included 18 metabolites (see Supplemental information for a complete list). Macromolecule signals included in the basis set were measured using inversion-recovery experiments [62] performed in the occipital cortex of 4 healthy adults [61]. Spectra were fitted using a relatively stiff baseline (spline constraint parameter DKNTMN = 5) without soft constraints on metabolite concentrations, as appropriate for these data [25,26,36]. We scaled our metabolite concentrations (millimolar) relative to water, and applied corrections for the particular gray matter (GM), white matter (WM), and cerebrospinal fluid (CSF) content within each VOI in each participant. The tissue fraction within each VOI in each participant was quantified using individual GM and WM surface models from FreeSurfer [version 5.3.0; 63]. See Supplemental information and Supplemental Table 1 for further details.

**Figure 2.**
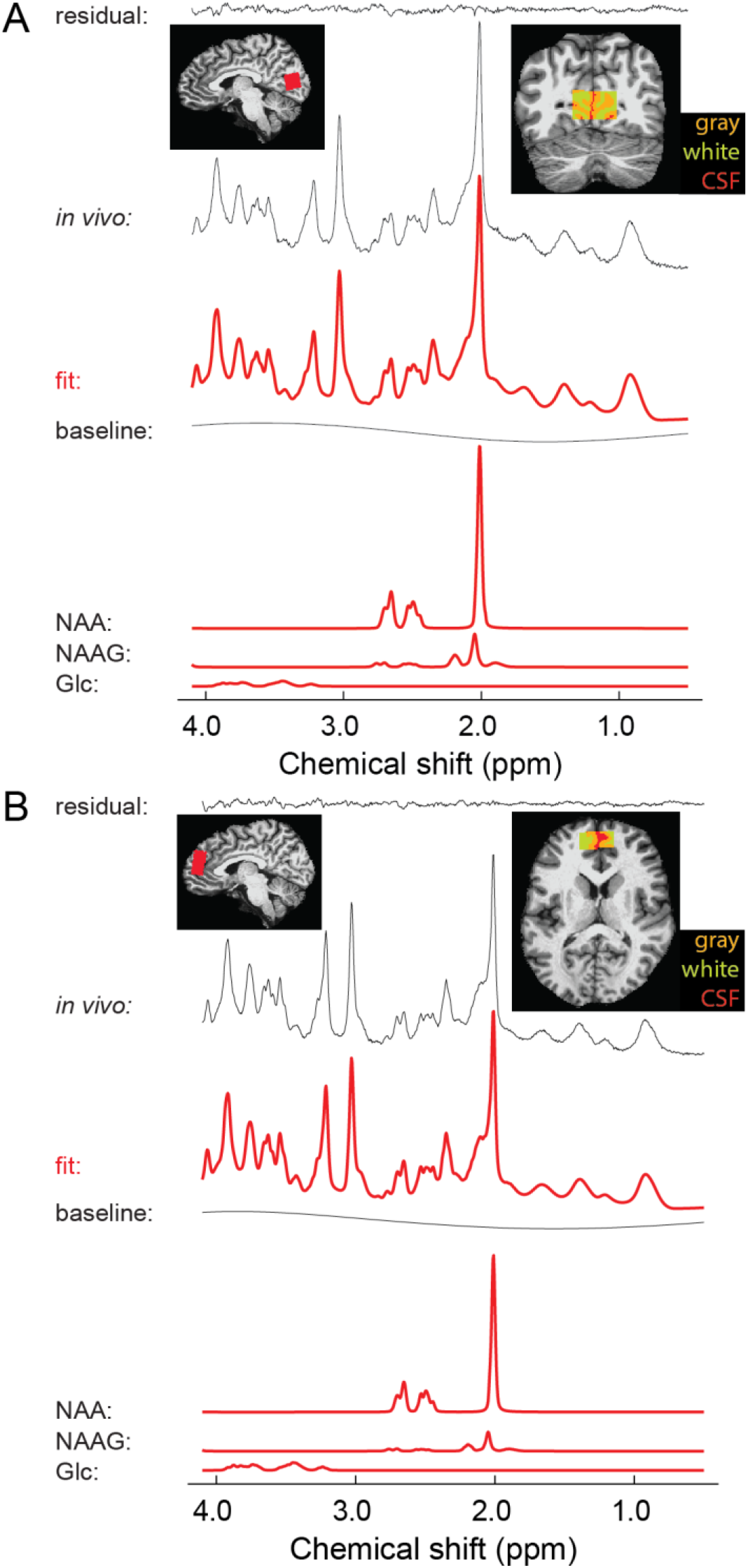
Example MR spectra. A) OCC spectrum from an individual participant (black) plus LCModel fit (red) and residual. The bottom rows show NAA, NAAG, and glucose (Glc) signals, as fitted by LCModel. Insets show OCC VOI position, top left: sagittal, with VOI in red; top right: coronal, with GM (orange), WM (yellow), and CSF (red) highlighted. B) Same, but for the PFC VOI; top right inset shows axial view.

We assessed data quality for OCC and PFC VOIs separately using metrics described in the Supplemental information. In total, 5 OCC data sets (2 relatives, 3 PwPP) and 9 PFC data sets (1 control, 5 relatives, 3 PwPP) were excluded due to poor quality. Please see [1] for additional visualizations and group comparisons of our MRS data quality metrics.

### Data analysis & statistics

We performed our analyses using MATLAB. We conducted two sets of analyses comparing metabolite concentrations from each VOI between groups. Our first analysis focused on 6 metabolites that were chosen *a priori*: GABA, glutamate, glutamine, glutathione, NAA, and NAAG. We chose these based on a review of the literature [11–17,64–66], which suggested that concentrations of these 6 metabolites in OCC and / or PFC may be altered in PwPP. As a number of our metabolite values were not normally distributed, and variance was not always equivalent across groups, we used Kruskal-Wallis 1-way non-parametric ANOVAs to assess group differences. We excluded repeat scan data, as Kruskal-Wallis tests assume all data points are independent. Because we considered the group comparisons for each of these 6 metabolites as separate hypotheses defined *a priori* based on the literature, we did not perform corrections for multiple comparisons in this case.

We also carried out a second set of *post hoc* group comparisons for 10 additional metabolites, and follow up analyses to examine the influence of covariates, as described in the Supplemental information.

A subset of participants participated in a repeat scanning session (Table 1). This allowed us to assess longitudinal variability for each of the 16 metabolites examined above. For this purpose, we quantified intra-class correlation coefficients (ICC, type (3,k); [67]). Note that of the 36 PwPP who completed repeat scanning sessions, OCC data from one or both sessions were excluded for 4 individuals. Likewise, PFC data were excluded for 2 PwPP who completed repeat sessions.

We performed hierarchical clustering analyses [68] (with *k* = 7 clusters) to examine multivariate relationships between metabolite concentrations from MRS, visual task performance metrics, and clinical measures in a data driven manner. This involved computing Spearman’s correlation values across all pairwise combinations of 30 variables (16 OCC metabolites, plus 6 visual task metrics, and 8 clinical measures; referred to as OCC clustering hereafter; Table 2) or 46 variables (the 30 above, plus 16 PFC metabolites; OCC+PFC clustering hereafter; Supplemental Table 2). We examined the statistical significance of our clustering results using permutation analyses. Additional details on these analyses are provided in the Supplemental information.

**Table 2.** List of variables for OCC hierarchical clustering analysis.

| Variable # | Variable type | Variable name |
| --- | --- | --- |
| 1 | Visual task | Contrast discrimination threshold |
| 2 | Visual task | Contrast surround suppression index |
| 3 | Visual task | Contour object discrimination accuracy |
| 4 | Visual task | Contour object jitter threshold |
| 5 | Visual task | Structure-from-motion bi-stable switch rate |
| 6 | Visual task | Structure-from-motion bi-stable coefficient of variance |
| 7 | Clinical measure | BPRS total score |
| 8 | Clinical measure | BACS Z-score |
| 9 | Clinical measure | SGI total score |
| 10 | Clinical measure | SPQ total score |
| 11 | Clinical measure | PID-5 psychoticism factor |
| 12 | Clinical measure | BPRS hallucination score |
| 13 | Clinical measure | Snellen visual acuity (decimal fraction) |
| 14 | Clinical measure | Mars visual contrast sensitivity |
| 15 | MRS measure | OCC ascorbic acid |
| 16 | MRS measure | OCC aspartic acid |
| 17 | MRS measure | OCC total creatine |
| 18 | MRS measure | OCC GABA |
| 19 | MRS measure | OCC glucose |
| 20 | MRS measure | OCC glutamine |
| 21 | MRS measure | OCC glutamate |
| 22 | MRS measure | OCC total choline |
| 23 | MRS measure | OCC glutathione |
| 24 | MRS measure | OCC *myo*-inositol |
| 25 | MRS measure | OCC macromolecules |
| 26 | MRS measure | OCC NAA |
| 27 | MRS measure | OCC NAAG |
| 28 | MRS measure | OCC phosphorylethanolamine |
| 29 | MRS measure | OCC *scyllo*-inositol |
| 30 | MRS measure | OCC taurine |

### Data and code availability

Our data are publicly available through the Human Connectome Project (db.humanconnectome.org) and the National Data Archive (nda.nih.gov/edit_collection.html?id=3162). Our data processing code is available from GitHub (github.com/mpschallmo/PsychosisHCP).

## Results

### Univariate results

We compared metabolite concentrations across our 3 groups (controls, relatives, PwPP) for the following 6 metabolites chosen *a priori*: GABA, glutamate, glutamine, glutathione, NAA, and NAAG. These comparisons were performed for both OCC and PFC VOIs separately.

In OCC, NAA levels differed significantly across groups (Kruskal-Wallis, *Χ*^2^_2, n = 141_ = 7.46, *p* = 0.024) and were lower for both PwPP and relatives compared to controls (post-hoc Kruskal-Wallis tests, *Χ*^2^ values > 4.35, *p* values < 0.037; Figure 3A). In PFC, we found a significant group difference in NAAG levels (*Χ*^2^_2, n = 112_ = 8.10, *p* = 0.017), which tended to be lower between PwPP compared to both relatives and controls (*Χ*^2^ values > 3.57, *p* values < 0.059; Figure 3B). Other *a priori* group comparisons for metabolites in OCC and PFC, including GABA and glutamate, were not significant (see Supplemental Figure 1 & Supplemental Figure 2, Supplemental Table 3).

**Figure 3.**
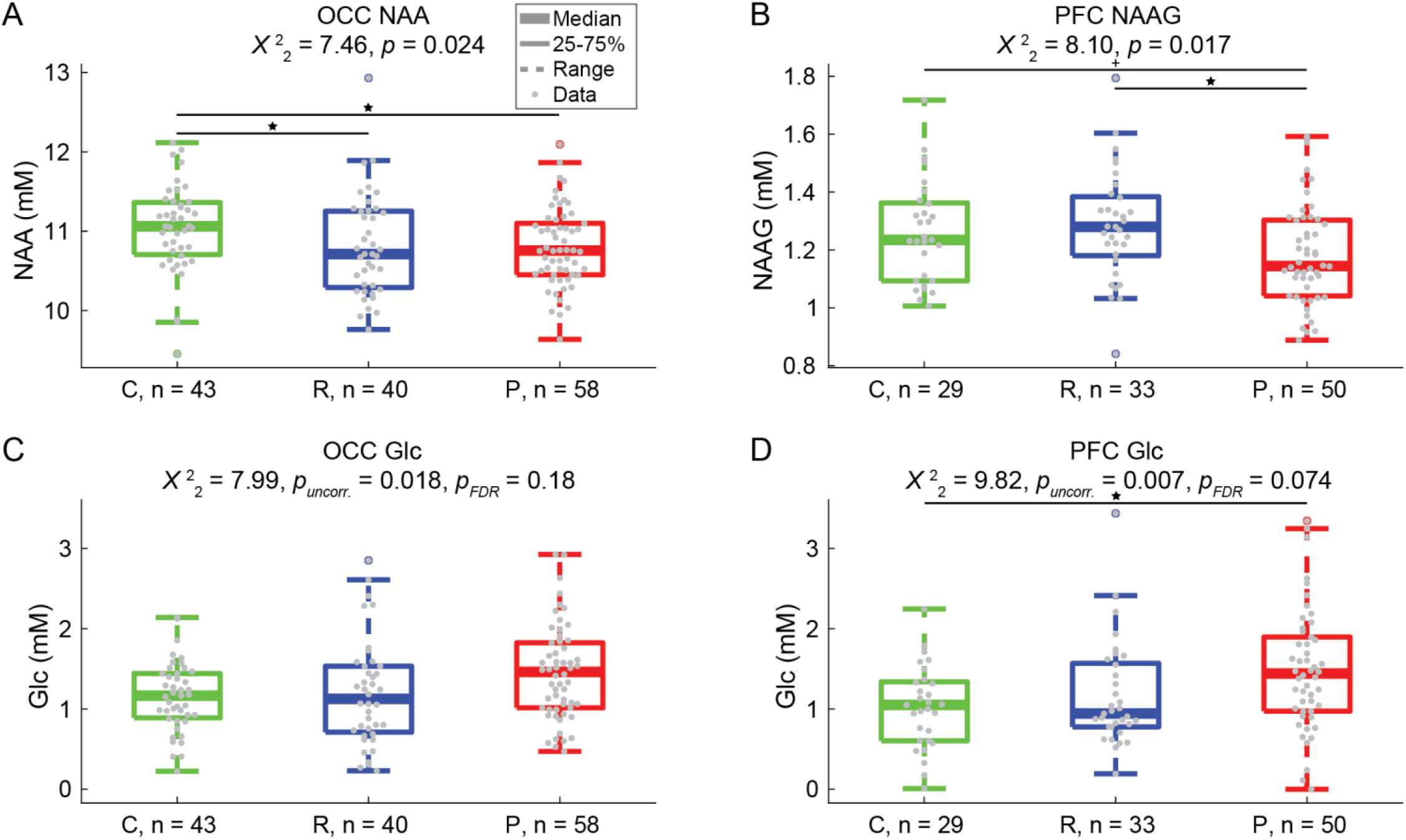
MRS results. We observed significant group differences in (A) NAA in OCC and (B) NAAG in PFC. Differences were also observed in glucose (Glc) in both OCC and PFC (C & D, respectively). C = controls, R = relatives, P = people with psychotic psychopathology. Thick lines show group medians, boxes show 25-75%, whiskers show 1.5x interquartile range, dots show individual data points. For comparisons across all 3 groups, *p* values in A & B are uncorrected, both uncorrected and FDR corrected *p* values are reported in C & D. Stars and lines indicate significant differences between 2 groups at *p* < 0.05 (after FDR correction in D), plus at *p* < 0.1. Repeat scan data are not shown.

Follow-up analyses with age, sex assigned at birth, and BMI as covariates suggested that the concentration of NAA in OCC was associated with age, but the difference in NAA between controls and PwPP (but not relatives) remained significant with these covariates (see Supplemental results). However, group differences in NAAG in PFC did not reach significance with age, sex, and BMI included as covariates. Neither the tissue composition within the VOIs, nor associations with antipsychotic medications, appeared sufficient to explain our findings of lower NAA in OCC and lower NAAG in PFC (see Supplemental results).

We also performed *post hoc* comparisons between groups for 10 additional metabolites in both OCC and PFC, with correction for multiple comparisons. Of these 10 metabolites, only glucose showed notable group differences. In PFC, there was a trend toward a group difference (*Χ*^2^_2, n = 112_ = 8.10, uncorrected *p* = 0.007, FDR-corrected *p* = 0.074), with significantly higher glucose levels in PwPP versus controls (*Χ*^2^_1, n = 79_ = 8.00, uncorrected *p* = 0.005, FDR-corrected *p* = 0.047; Figure 3D), and relatives showing intermediate values (relatives vs. PwPP: *Χ*^2^_1, n = 83_ = 5.50, uncorrected *p* = 0.019, FDR-corrected *p* = 0.19). A similar pattern was observed in OCC, but was not significant after correcting for multiple comparisons (*Χ*^2^_2, n = 141_ = 7.99, uncorrected *p* = 0.018, FDR-corrected *p* = 0.18; Figure 3C). For data on other metabolites, see Supplemental Figure 1 & Supplemental Figure 2, and Supplemental Table 3. Thus, our MRS analyses revealed unexpectedly higher glucose levels in PwPP.

We asked whether demographic differences between groups (e.g., age, BMI; Table 1) might account for our observation of higher glucose levels between PwPP and relatives. Because glucose is present in CSF and can be measured using MRS [69–71], we also asked whether group differences in CSF fraction may have influenced our results. However, follow-up analyses suggested that factors such as age, sex, BMI, antipsychotic medication levels, and tissue composition within the VOI may not have been sufficient to fully account for the group differences we observed in glucose (see Supplemental results).

Finally, we examined differences in metabolite levels between people in the PwPP group with diagnoses of schizophrenia versus bipolar disorder, as a complement to our trans-diagnostic analyses above. People with schizoaffective disorder (n = 9) were excluded from this analysis, due to low sample size. We compared the 6 metabolites that we had chosen to focus on *a priori*, in addition to glucose, in both OCC and PFC. None of these differences were significant (*Χ*^2^ values < 3.4, uncorrected *p* values > 0.067; data not shown).

### Longitudinal variability

A subset of participants (10 controls, 36 PwPP) returned for a repeat scanning session in which MRS data were acquired from the same VOIs in OCC and / or PFC. This allowed us to assess longitudinal variability in metabolite concentrations over a period of several months (see Table 1 for information about the time between scanning sessions). VOI placement tended to overlap highly across sessions (see Supplemental results).

Agreement between the two sessions ranged from fair to excellent across all 16 metabolites (range of ICC values: 0.44 to 0.96; Figure 4; Supplemental Table 4). These results indicate that metabolite concentrations were relatively stable within individuals across time in our sample.

**Figure 4.**
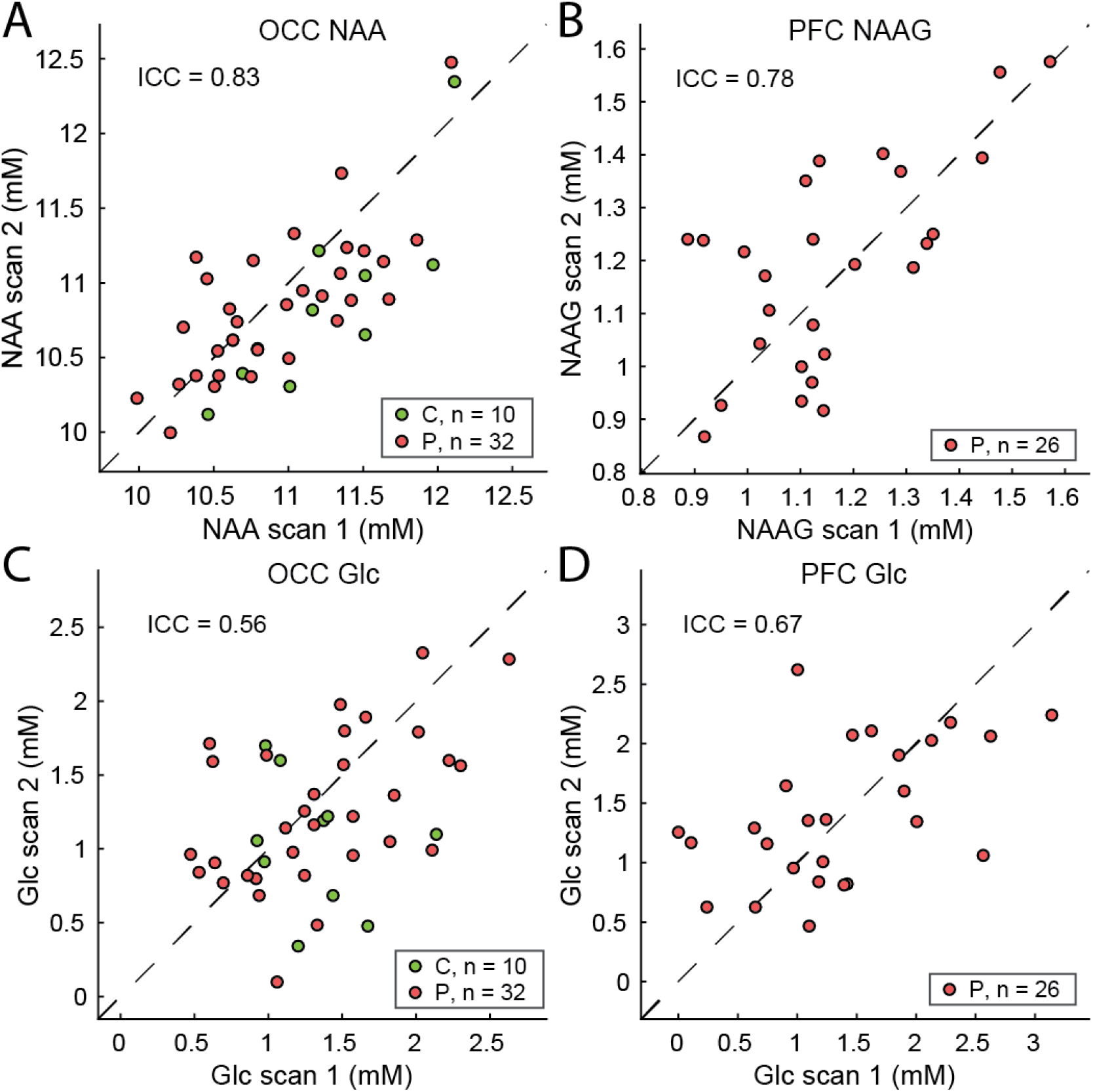
Longitudinal variability of MRS data. Variability in metabolite concentrations was quantified across 2 scanning sessions within a subset of participants (10 controls, C, green; 36 PwPP, red). A) NAA levels in OCC, B) NAAG levels in PFC, C) glucose (Glc) levels in OCC, D) Glc levels in PFC. Intra-class correlation coefficients (ICC) across all participants are shown in each panel. Dashed black lines show unity. Note that of the 36 PwPP who completed repeat scanning sessions, OCC data from one or both sessions were excluded for 4 individuals. Likewise, PFC data were excluded for 2 PwPP who completed repeat sessions.

### Hierarchical clustering

These MRS data were acquired as part of a larger set of experiments within the Psychosis Human Connectome Project [1,43], which included behavioral measures of visual task performance and clinical assessments of psychiatric functioning (see Methods, Supplemental Information). We performed a hierarchical clustering analysis in order to understand the pattern of multivariate relationships between our metabolite levels from MRS and these visual and clinical measures. This involved calculating pairwise correlations between all variables (listed in Table 2), and then quantifying which groups of variables were most strongly correlated with one another (see Methods, Supplemental information).

Our OCC clustering analysis showed that glucose in OCC (variable #19, red arrow in Figure 5) was part of a cluster of variables (red dashed box in Figure 5) that included psychiatric measures (e.g., Brief Psychiatric Rating Scale total score [#7], Brief Assessment of Cognition in Schizophrenia [#8]), as well as measures of visual task performance (i.e., contrast discrimination thresholds [#1], bi-stable illusion switch rates [#5]; see Table 2). Other MRS measures (#15-30) tended to cluster with one another (yellow and blue dashed boxes in Figure 5) and not with the visual and clinical measures. A dendrogram illustrating the OCC clustering results is presented in Supplemental Figure 3. We performed permutation analyses to assess the statistical significance of our hierarchical clustering results. For the cluster that included glucose in OCC (red dashed box in Figure 5), the average *r* value was significantly higher than values obtained using shuffled data (permutation test with data shuffled across participants: *p* < 0.001; with variables shuffled across clusters: *p* = 0.022).

**Figure 5.**
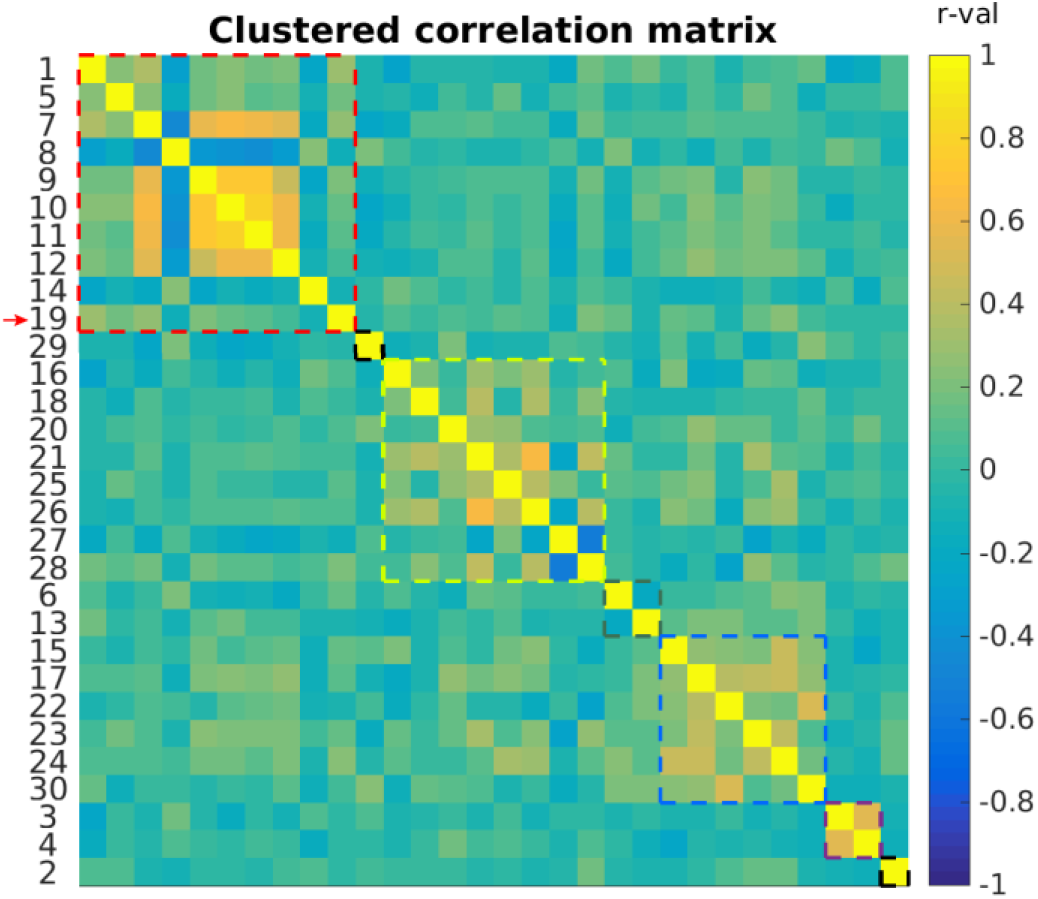
OCC hierarchical clustering results. Cells show pairwise Spearman correlation *r* values for measures from visual tasks (variables #1-6), clinical assessments (#7-14), and MRS in OCC (#15-30). See Table 2 for a full list of variables by number. Colored dashed lines indicate the boundaries of various clusters (*k* = 7). Red arrow indicates glucose (#19), which was the only MRS variable that tended to cluster with visual task and clinical measures (red dashed box).

We observed similar results for OCC+PFC clustering (Supplemental Figure 4 & Supplemental Figure 5). Glucose in both OCC and PFC (variables #19 & 35) tended to cluster with the same psychiatric measures and visual task variables as in Figure 5. Interestingly, *scyllo*-inositol levels from both OCC and PFC (variables #29 & 45) were also included within this cluster.

Permutation analyses again showed that the average *r* value within the cluster that included glucose levels in OCC and PFC was significantly higher than for shuffled data (permutation test with data shuffled across participants: *p* = 0.002; with variables shuffled across clusters: *p* = 0.041). Together, the results of our hierarchical clustering analyses indicate that across participants, higher glucose levels in OCC and PFC are associated with higher psychiatric symptom levels, and greater abnormalities in visual task performance.

## Discussion

Using data we collected as part of the Psychosis HCP, we examined differences in the concentrations of 16 neurochemicals between PwPP, biological relatives, and controls in both OCC and PFC. Among the 6 metabolites we examined *a priori*, we found lower NAA levels in OCC in PwPP and relatives, and lower NAAG in PFC in PwPP, which agrees in part with previous studies [16,64,72]. Our *post hoc* analyses also revealed significantly higher glucose levels in PwPP in PFC, and a similar trend in OCC. A subset of our participants returned for a repeat scanning session, and our metabolite measurements tended to be reliable across time. Together, our results reveal a pattern of metabolic dysfunction across brain regions in PwPP, and to a lesser extent in their biological relatives.

In our *a priori* univariate analyses, we found that NAA levels in OCC were significantly lower in both PwPP and relatives, as compared with controls. We used an ultra-short (8 ms) TE sequence in order to address concerns that *T_2_* decay rates for some metabolites, including NAA, might be shorter in PwPP [27–30]. Our observation that OCC NAA levels are lower in PwPP and their relatives suggests that there may be a true reduction in occipital NAA in these groups, beyond a possible reduction in *T_2_*. Our finding also suggests that reductions in occipital NAA may be linked to genetic liability for psychosis, as expressed by first-degree biological relatives, who share on average 50% of their genes with a person with a psychotic disorder. However, as age was associated with lower NAA levels, we cannot rule out the possibility that lower NAA levels in relatives may have been driven by the fact that this group tended to be older on average compared to controls.

NAA plays multiple roles in the human brain, which include providing storage for acetyl units that are used in myelin synthesis, regulating osmotic pressure, and supporting neural metabolic functions [73]. NAA levels are thought to reflect the level of neuronal metabolic activity and / or neuronal density within a given brain region [73]. As NAA synthesis is coupled to glucose metabolism [74], our observations of higher glucose and lower NAA levels in OCC may both reflect metabolic dysfunction in PwPP. Reduced occipital NAA has been observed previously in people with psychotic disorders including schizophrenia [16]. Our study builds upon these findings by showing this deficit is present in a relatively large, trans-diagnostic group of PwPP, as well as their biological relatives, assessed at 7 T. To date, comparatively few studies have examined NAA levels in biological relatives of PwPP; these have generally shown reduced NAA concentrations for relatives versus controls across several brain regions [18–20,22].

NAA is converted to NAAG through the addition of glutamate. NAAG is a peptide neurotransmitter that is thought to act as an agonist at metabotropic glutamate receptors (i.e., mGluR2/3 [75,76], but see [77] for conflicting results). The ability to reliably quantify NAAG using MRS is hampered by relatively low concentrations in the human brain, and spectral overlap with other metabolites present at higher concentrations, specifically NAA [23]. Often, these two metabolites are quantified together (NAA+NAAG). Here, separate quantification was facilitated by our use of 7 T MRS, and our high spectral quality (in terms of linewidth). Although the precise roles of NAAG as a neurotransmitter remain under investigation, differences in NAAG levels have been observed in schizophrenia [64,72]. In the current study, we found lower NAAG levels in PwPP as compared to relatives and controls in PFC, but no difference in OCC. However, we found that this difference was no longer significant after demographic factors (e.g., age) were included as covariates (see Supplemental results). Although higher NAAG levels in PFC correlated with a higher proportion of white matter within the voxel, we saw no group differences in tissue composition (see Supplemental results). In general, the observation of lower NAAG levels in PFC in PwPP appears broadly consistent with the idea of abnormal glutamatergic functioning in this population [3,78].

It is unclear to what extent lower NAA levels in OCC and NAAG levels in PFC may reflect a common anomaly in PwPP, or may instead be separate deficits. NAA is a precursor for NAAG; these two metabolites were modestly but significantly anti-correlated in OCC (*r*_147_ = −0.23, *p* = 0.005; variables #26 & 27 in Figure 5, respectively), and to a lesser extent in PFC (*r*_108_ = −0.18, *p* = 0.066; variables #42 & 43 Supplemental Figure 4). In our hierarchical clustering analysis, NAA and NAAG in OCC and PFC tended to cluster together (light blue box in Supplemental Figure 4), consistent with the notion that reductions in these metabolites may reflect one common source of metabolic dysfunction. On the other hand, NAA in OCC was lower in relatives vs. controls, whereas NAAG in PFC was normal in relatives. This suggests that group differences in NAA vs. NAAG are to some extent distinct, may vary across brain regions, and that NAA in OCC may reflect genetic liability for psychotic disorders (as present in biological relatives), whereas NAAG levels in PFC may be more closely linked to disease processes in PwPP.

We observed higher glucose levels in PwPP in PFC, and to a lesser extent in OCC, in our *post hoc* analyses. Our hierarchical clustering results indicated that glucose levels in OCC and PFC were part of a group of associated variables that also included measures of clinical psychiatric symptoms and visual task performance. The association between these variables may reflect a single latent construct underlying aspects of cognitive and perceptual dysfunction in people with psychosis, such that PwPP who have higher concentrations of brain glucose also experience greater perceptual and cognitive impairments. Impaired brain metabolism among PwPP has also been found in studies using other imaging modalities, including reports of lower glucose metabolism as observed by ^18^FDG-PET, especially in the frontal lobe in people with schizophrenia [10]. Our observation of higher glucose levels in PFC via ^1^H-MRS appears broadly consistent with the notion of hypofrontality in PwPP, which may contribute to psychiatric symptoms. Higher glucose levels may reflect metabolic dysfunction (i.e., reduced glucose utilization in the brain) in PwPP *per se*, or may be a secondary effect of medication. In particular, metabolic side effects including weight gain, increased blood glucose, type 2 diabetes mellitus are common in individuals taking second generation antipsychotic medications [79,80]. In the current study, we found that the level of glucose in OCC (but not PFC) was positively correlated with BMI across all participants (but not within the relative or PwPP groups). We also found no significant associations between brain glucose and antipsychotic medication levels in our sample.

We did not expect to find higher glucose levels in PwPP based on the existing MRS literature at the inception of our study in 2017. However, similar results have recently been reported by Wijtenburg and colleagues [9]. Using an ultra-short echo time (6.5 ms) STEAM sequence at 3 T, they found higher glucose levels in medial occipital cortex in a modest number of people with schizophrenia (n = 16) versus controls (n = 19). Increased occipital glucose was correlated with poorer performance on the Brief Visuospatial Memory Test, and abnormal insulin resistance biomarker levels in their study. Thus, our observation that higher brain glucose was associated with impairment in a cluster of psychiatric, cognitive, and visual task performance measures appears consistent with these recent findings [9]. Detection of glucose via MRS is complicated by overlap with resonances from other metabolites, and relatively low concentrations of glucose in the brain (∼ 1 mM) [23].

Both our study and that of Wijtenburg and colleagues [9] used ultra-short TE (≤ 8 ms) STEAM sequences, which may facilitate quantification of glucose by minimizing signal loss due to *T_2_* decay and phase dispersion from *J* evolution. For this reason, it is possible that other previous MRS studies using sequences with longer TEs may have failed to detect higher levels of brain glucose between PwPP and controls.

Our study has a number of strengths, including a relatively large sample size for a study of PwPP conducted at a single site. Conducting our experiments at 7 T gave us higher SNR and improved our ability to resolve individual metabolites (e.g., separately quantifying glutamate vs. glutamine, reliably quantifying GABA), as compared to MRS methods at 3 T [31–35]. We explicitly accounted for macromolecules in our basis set using the results from inversion recovery experiments conducted in a previous study [61]. Further, we fitted our data using a rigid baseline and without implementing soft constraints on metabolite values, as appropriate for high-quality, artifact-free spectra. Recent work has shown that the use of soft constraints and / or more flexible baseline settings, which are the default for LCModel, can lead to differences in metabolite quantification as compared to a fully specified modeling approach [25,26,36]. We also used an ultra-short TE (8 ms) STEAM sequence to address concerns about shorter *T_2_* relaxation time constants in people with psychosis [27–30].

The above methodological factors may help to explain why we observed group differences for some, but not all, of the metabolites we examined *a priori*. For example, group differences in glutamate or GABA levels reported in previous studies might in fact reflect differences due to quantification errors and / or shorter *T_2_* relaxation time constants in PwPP. Other strengths included the fact that we included biological relatives in the current study, which allowed us to examine the role of genetic risk for psychosis in our brain metabolite measures. Because there are concerns regarding the validity and reliability of traditional psychiatric diagnostic categories [41,42], we chose to adopt a transdiagnostic approach to studying neurochemistry in PwPP. Thus, the differences in brain chemistry we observed may be more broadly reflective of psychotic psychopathology, instead of a particular diagnosis (e.g., schizophrenia). However, we acknowledge that this approach necessarily limited our ability to make comparisons between diagnostic groups, due to limitations on sample size. Another limitation was that only a subset of participants (10 controls, 39 PwPP) completed a repeat scanning session. Thus, our ability to examine longitudinal variation of brain metabolite levels within individuals was limited. Finally, our sample was limited to individuals who could fit comfortably in the scanner, and who has successfully completed two prior scanning sessions at 3 T. Given our results, further studies using MRS to understand the role of excess brain glucose among PwPP appear warranted.

## Data Availability

Our data are publicly available through the Human Connectome Project and the National Data Archive.

https://db.humanconnectome.org

https://nda.nih.gov/edit_collection.html?id=3162

## Acknowledgments

We thank Jesslyn (Li Shen) Chong, Victoria Espensen-Sturges, Andrea N. Grant, Rohit S. Kamath, Samantha A. Montoya, Hannah R. Moser, Marisa J. Sanchez, Karina Smiley, and Kimberly B. Weldon for their assistance with data collection and / or processing. We also thank Edward J. Auerbach for implementation of STEAM and FAST(EST)MAP sequences on Siemens scanners, Dinesh K. Deelchand for simulation of the basis set, and Guglielmo Genovese for code used in generating Figure 2.

This work was supported by funding from the National Institutes of Health (U01 MH108150). Salary support for MPS was provided in part by K01 MH120278. Support for MR scanning at the University of Minnesota Center for Magnetic Resonance Research was provided by P41 EB015894 and P30 NS076408. This work used tools from the University of Minnesota Clinical and Translational Science Institute that were supported by UL1 TR002494.

## Supplemental information

### Supplemental methods

#### Participants

Participants were 18-65 years old, spoke English as a primary language, and did not have: a diagnosed learning disability or IQ less than 70, current or past central nervous system disease, history of head injury with skull fracture or loss of consciousness longer than 30 minutes, alcohol or drug abuse within the last 2 weeks, or dependence within the last 6 months, electroconvulsive therapy within the last year, tardive dyskinesia, visual or hearing impairment. All participants were able to fit into the MRI scanner comfortably and had a corrected Snellen visual acuity of 20/40 (decimal fraction = 0.5) or better. All participants in the PwPP group had a history of schizophrenia, schizoaffective disorder, or bipolar I disorder with psychotic features. Relatives had a biological parent, sibling, or child with a history of one of these disorders. Relatives included people with and without current psychiatric diagnoses (Table 1). Controls had no personal or immediate family history of psychosis spectrum disorders. We assessed each participant’s capacity to provide informed consent using the University of California Brief Assessment of Capacity to Consent [81].

#### Apparatus

Scanner software versions were VB17 (before July, 2019) and VE12U (after July, 2019). Participants were given head, neck, and lumbar padding to increase their comfort during scanning. We used an in-scanner motion tracking system from Metria Innovation Inc., (Milwaukee, WI) with a Moiré Phase Tracking marker attached to the participant’s nose to monitor head motion during scanning. Experimenters monitored head motion in real time during the scan, and when motion > 5 mm was observed, MRS scans were repeated and participants were encouraged to remain still.

#### MR spectroscopy

STEAM sequence parameters were as follows: repetition time (TR) = 5 s, echo time (TE) = 8 ms, mixing time (TM) = 32 ms, hsinc pulse = 1.28 ms, transmitter frequency = 3 parts per million (ppm), number of complex data points = 2048, spectral bandwidth = 6000 Hz, chemical shift displacement error = 4%. We used 3D outer volume suppression interleaved with variable power and optimized relaxation delay (VAPOR) water suppression [82]. We adjusted the transmit voltage for each VOI in order to calibrate the B_1_ field for the 90° pulse and the water suppression flip angles.

We collected 128 shots in the PFC VOI for an initial group of 30 participants to assess data quality. After confirming that our PFC data were of comparable quality to those from our OCC VOI, the number of shots was reduced to 96 for subsequent participants. Our frontal MRS coil was not available during the first 6 months of data collection, which meant that we were not able to collect PFC data from all participants. OCC data collection always preceded PFC. Participants were removed from the scanner between OCC and PFC scans to switch from occipital to frontal transceiver coils.

We acquired a *T*_1_-weighted anatomical scan during our MRS experiment, which we used to guide voxel placement. *T*_1_ scan parameters were as follows: TR = 2500 ms, TE = 3.2 ms, echo spacing = 7.9 ms, flip angle = 5°, resolution = 1.3 mm isotropic, partial Fourier = 6/8, 64 mid-sagittal slices, field of view = 160 x 160 mm^2^.

#### Visual tasks

We collected data from the visual tasks described below as part of the Psychosis Human Connectome Project. For full details, please see our recent publication [1]. Data were collected outside the scanner, prior to MR scanning, typically as part of the same study visit. Tasks were performed using PsychoPy [83] on a Mac Pro with a linearized FlexScan SX2462W monitor (60 Hz refresh rate, mean luminance = 61.2 cd/m^2^, viewing distance 70 cm) and a Bits# stimulus processor.

The first visual paradigm was designed to measure contrast discrimination thresholds for both isolated grating stimuli, as well as gratings presented in the context of a high contrast surrounding annular grating [84–88]. The former provided a measure of contrast sensitivity, whereas the latter measured the modulation of contrast discrimination by the surrounding grating (i.e., surround suppression). Eight stimulus conditions were used.

The first seven included target gratings without surrounds at 0, 0.6, 1.25, 2.5, 5, 10, and 20% pedestal contrast. The eighth used 10% contrast targets with 100% contrast surrounds. Targets and surrounds (when present) were presented on both sides of a central fixation mark at 3° eccentricity (target diameter = 2°, surround inner and outer diameter = 2.5° and 4°, respectively, spatial frequency = 1.1 cycles/°, contrast reversing at 4 Hz). On each trial, targets appeared for 800 ms, followed by an unlimited response period. A contrast increment was added to the target grating on one side (randomized) on every trial. The participants’ task was to indicate which target was higher contrast (left or right). The increment value was controlled by a Psi adaptive staircase across trials [89]. This allowed us to find an increment value at which the participants could reliably discriminate which target was higher contrast. Each staircase included 30 trials, and there were 3 independent staircases per stimulus condition. Conditions were presented in separate blocks. Threshold values were calculated at 80% accuracy by fitting the staircase data for each condition with a Weibull function. For this study, we calculated the average contrast discrimination threshold (variable #1, Table 2 & Supplemental Table 2) by taking the geometric mean of the threshold values from the 7 conditions without surrounds. We calculated a contrast surround suppression index (variable #2, Table 2 & Supplemental Table 2) by taking the ratio of the thresholds for targets at 10% contrast with surrounds / targets without surrounds (at 10% contrast).

The second visual task focused on contour object perception. Discrete visual elements (i.e., Gabors, *SD* = 0.067°, spatial frequency = 5 cycles/°) composed an egg shaped contour (15 elements, egg was 5.9° wide x 4.7° high). Participants had to perceptually integrate the contour elements in order to discriminate the directionality of the egg shape (i.e., pointing left or right) within a field of 170 randomly oriented background distractor elements [90–93]. Data from two types of contour object perception trials were used in the current study. In the first type, the orientation of the contour elements was perfectly aligned with the axis of the egg shaped contour. In the second type of trials, orientation jitter was added such that the Gabors that composed the contour were miss-aligned with the contour axis; this was done in order to make the directionality of the contour more difficult to perceive. Jitter was controlled across trials using a Psi adaptive staircase, in order to find the jitter level at which participants could reliably discriminate left-versus right-facing contours. Staircases for the jittered contour condition included 30 trials, and there were 3 independent staircases in each of two block (6 total staircases). Jitter threshold values at 70% accuracy were obtained by fitting the staircase data from each block with a Weibull function. There were also 20 trials of the aligned contour condition in each block (40 total). For this study, we calculated the average accuracy for contour object discrimination (variable #3, Table 2 & Supplemental Table 2) across trials in the aligned contour condition. We also calculated the average jitter threshold (variable #4, Table 2 & Supplemental Table 2) across the two blocks.

Our third visual task focused on a bi-stable visual structure-from-motion paradigm known as the rotating cylinder illusion [94,95]. In this task, participants viewed an array of 400 squares (0.25° wide, 200 black, 200 white) that moved horizontally across the screen in a manner that simulated the rotation of a 3 dimensional cylinder in depth (height = 10°, width = 7°, rotation speed = 90°/s). Because the stimulus lacked depth cues, the direction of rotation (i.e., clockwise or counter-clockwise in depth) was ambiguous, which caused the image to appear to alternate between these two perceptual interpretations spontaneously in a bi-stable manner.

Participants reported their initial percept and all subsequent perceptual switches by pressing a key to indicate which direction the front face of the cylinder appeared to rotate. Participants viewed and responded to a continuously rotating image for 2 minutes in each of 5 task blocks. Based on the timing of reported perceptual switches, we calculated the average structure-from-motion bi-stable switch rate across blocks (variable #5, Table 2 & Supplemental Table 2), as well as the coefficient of variance (i.e., the standard deviation of the percept durations, divided by the mean; variable #6, Table 2 & Supplemental Table 2).

All three of our visual tasks included catch trials in order to assess the participants’ ability to understand and carry out the task as instructed. Data from participants who failed to achieve sufficiently high performance in these catch trials were excluded; see our recent paper for full details [1].

#### Clinical measures

Clinical measures included questionnaires and interview-based instruments relevant to psychotic psychopathology. For full details, please see our recent publications [1,43]. This included the Brief Psychiatric Rating Scale (BPRS) [48]; this interview-based measure focused on psychiatric status over the past 30 days, and was typically collected during the same experimental session as the 7T MRS scan (but always within 30 days prior to scanning). For this study, we used both the BPRS total score (variable #7, Table 2 & Supplemental Table 2) and the score for the hallucinations item (variable #12). We chose to include the hallucination score in particular due to our specific interest in sensory dysfunction in PwPP. We also assessed visual acuity using a Snellen eye chart (computed as a decimal fraction, e.g., 20/40 = 0.5; variable #13, Table 2 & Supplemental Table 2) [44] during these 7T sessions. Other measures were collected at a separate study visit prior to MR scanning, and included the Brief Assessment of Cognition in Schizophrenia (BACS; *Z*-scored, variable #8, Table 2 & Supplemental Table 2) [47], the Sensory Gating Inventory (SGI; total score, variable #9) [49], the Schizotypal Personality Questionnaire (SPQ; total score, variable #10) [50], the Personality Inventory for the DSM-5 (PID-5; psychoticism factor score; variable #11) [51], and the Mars Letter Contrast Sensitivity test (variable #14) [45].

#### Data processing

We processed our data using the *matspec* toolbox (<u>github.com/romainVala/matspec</u>) in MATLAB. In the rare cases where we found that frequency and / or phase correction failed (based on visual inspection), we manually excluded individual shots. We also manually excluded a small number of full data sets (OCC = 3, PFC = 5) with significant artifacts (e.g., failed water suppression, defined as a water peak > NAA peak). We used a basis set from a previous study [61], which included 18 metabolites: ascorbic acid, aspartic acid, creatine, γ-aminobutyric acid (GABA), glucose, glutamate, glutamine, glutathione, glycerophosphorylcholine, lactate, *myo*-inositol, *N*-acetyl-aspartate (NAA), *N*-acetyl-aspartylglutamate (NAAG), phosphocreatine, phosphorylcholine, phosphorylethanolamine, *scyllo*-inositol, taurine, in addition to lipids and macromolecules.

Tissue fractions (gray matter, white matter, and CSF) were obtained from *T*_1_- and *T*_2_-weighted anatomical scans acquired at 3 T in a separate scanning session [43], and then processed through the Human Connectome Project minimal preprocessing pipeline [version 3.22.0; 96]. To obtain tissue fraction values for each MRS VOI, we aligned the partial sagittal *T*_1_-weighted anatomical data from the MRS session to the whole brain *T*_1_ scan acquired at 3 T. Water scaling correction values are reported in Supplemental Table 1. We did not correct for *T*_1_ and *T*_2_ relaxation times for our metabolites, as these effects are expected to be very small given the long TR (5 s) and ultra-short TE (8 ms).

We used following three MRS data quality metrics. First, we set a maximum value for water linewidth of 15 Hz, to ensure sufficient shim quality and *B*_0_ homogeneity within our VOIs. This value was determined *a priori*, and was assessed during each scan using tools from the *Spectroscopy* tab in the Siemens console software. In general, if we obtained a water linewidth > 15 Hz, the VOI was repositioned and re-shimmed. However, in a small number of cases (n = 3), MRS data were acquired with a water linewidth > 15 Hz. We also used two *post hoc* data quality metrics provided by LCModel. Based on visual inspection of our LCModel data, we set maximum values for spectrum linewidth = 5 Hz and SNR = 40. LCModel quantifies SNR based on the maximum signal value (i.e., from the NAA peak) divided by two times the root mean squared value of the residuals. Data sets that failed to meet one or more of these quality metrics were excluded.

#### Data analysis and statistics

Our *post hoc* analyses included the following 10 metabolites: ascorbic acid, aspartic acid, total choline, total creatine, glucose, *myo*-inositol, phosphorylethanolamine, *scyllo*-inositol, taurine, and macromolecules. We did not examine group differences for lactate or lipids, as they were not fit consistently across all participants. Total choline was calculated as the sum of the fit glycerophosphorylcholine and phosphorylcholine values, whereas total creatine was calculated from the sum of the fit creatine and phosphocreatine values. Here, we performed false discovery rate (FDR) correction for multiple comparisons (10 tests within each VOI).

Our supplemental analyses examined whether demographic or other factors might account for the observed group differences in metabolite levels reported in the main text. These analyses were performed in MATLAB. For NAA in OCC and NAAG in PFC, we used linear mixed effects models to assess group differences in metabolite levels, with age, sex, and BMI included as covariates. For glucose in OCC and PFC, variance was not equal between groups based on visual inspection (Figure 3C & D). Therefore, instead of using linear mixed effects models, we examined differences in glucose levels between individuals assigned male and female at birth using Kruskal-Wallis 1-way non-parametric ANOVAs, as well as Spearman correlations between glucose and both age and body mass index (BMI).

To assess whether tissue composition within our VOIs may have been associated with group differences in metabolite levels, we calculated the proportion of gray matter (GM), normalized by the summed proportions of gray matter and white matter (WM), as follows:

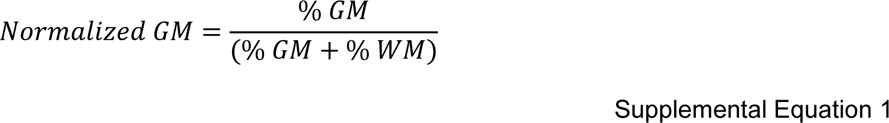

The proportions of GM, WM, and CSF within each VOI were obtained from FreeSurfer [63], as described in the Methods. We assessed group differences in normalized GM and CSF fractions using Kruskal-Wallis 1-way non-parametric ANOVAs. We also examined Spearman correlations between these tissue composition metrics and metabolite levels (NAA and glucose in OCC, NAAG and glucose in PFC).

We also examined whether antipsychotic medication levels among PwPP may have been associated with metabolite concentrations. For this purpose, we quantified chlorpromazine (CPZ) dose equivalents using established methods [54,55]. We used Spearman correlations to test associations between CPZ equivalents and NAA and glucose in OCC, as well as NAAG and glucose in PFC.

We used Dice coefficients to calculate the overlap in VOI position within participants across repeated scanning sessions. The Dice coefficient is a value that ranges between zero and one, and reflects the degree of overlap in 3D space between two binary data sets. We calculated Dice coefficients between binary VOI masks aligned to the same FreeSurfer [63] *T*_1_ anatomical reference scan using AFNI’s *3ddot* function [97].

For our hierarchical clustering analysis [68], data from repeat scanning sessions were excluded, as Spearman’s correlation method assumes independent data points. We calculated a similarity measure for this analysis, which we refer to as absolute correlation distance. This was quantified as one minus the absolute value of the Spearman correlation coefficients. Cluster analysis was performed with the *linkage.m* and *cluster.m* functions in MATLAB. Linkage was computed using the unweighted average distance. Because there is no formal rule for determining the number of clusters in this kind of hierarchical analysis, we examined the patterns of results when varying the number of clusters (*k*) from 2 to 15 for both our OCC and OCC+PFC clustering analyses (see dendrograms in Supplemental Figure 3 & Supplemental Figure 5). Because the results, as shown in the dendrograms, did not reveal an obvious stopping point for clustering, we chose to focus on the results obtained with *k* = 7 clusters, as they were broadly representative of the observed pattern of results as a whole. To assess the statistical significance of our clustering results, we performed two permutation analyses: 1) with data randomly shuffled across participants prior to clustering (to generate a null distribution of *r* values), and 2) with variables randomly shuffled across clusters (to generate a null distribution of clusters based on observed *r* values). For both analyses, we calculated the mean of all *r* values within each cluster (*k* = 7) after taking the absolute value. This was repeated across 1000 iterations. We quantified significance (i.e., *p* value) based on the proportion of permuted *r* values generated under the null distribution that were larger than the observed mean absolute cluster *r* values.

### Supplemental results

#### Demographics

Because our groups differed on demographic factors such as age and BMI (see Table 1), we performed the following additional analyses to examine whether these factors may have influenced the results of our univariate analyses reported in the main text. We compared group differences in metabolite levels using linear mixed-effects models with age, sex, and BMI included as covariates. We found that NAA levels in OCC were significantly associated with age (linear mixed-effects model, *t*_156_ = −3.38 parameter estimate [SE] = −0.014 [0.004], *p* = 9 x 10^-4^), but not sex assigned at birth (*t*_156_ = −1.20, parameter estimate [SE] = −0.11 [0.09], *p* = 0.2) or BMI (*t*_156_ = −0.32, parameter estimate [SE] = −0.003 [0.009], *p* = 0.7). With these covariates included, NAA was significantly lower in PwPP compared with controls (*t*_156_ = −2.12, parameter estimate [SE] = −0.24 [0.11], *p* = 0.035). However, the difference between controls and relatives was not significant (*t*_156_ = −1.24, parameter estimate [SE] = −0.16 [0.13], *p* = 0.2). Thus, we cannot rule out the possibility that lower NAA levels in OCC in relatives in particular may have been driven by the fact that this group was older than the controls on average.

We performed the same analyses for NAAG levels in PFC, which were also significantly associated with age (linear mixed-effects model, *t*_113_ = 4.43, parameter estimate [SE] = 0.006 [0.001], *p* = 2 x 10^-5^), but not sex assigned at birth (*t*_113_ = 1.13, parameter estimate [SE] = 0.038 [0.033], *p* = 0.2) or BMI (*t*_113_ = 0.53, parameter estimate [SE] = 0.002 [0.003], *p* = 0.6). However, after accounting for these variables, the difference in NAAG between PwPP and controls was not significant (*t*_113_ = 1.45, parameter estimate [SE] = 0.002 [0.042], *p* = 0.13). Thus, we cannot exclude demographic differences as a potential explanation for our observation of lower NAAG in PFC in PwPP.

We did not use linear mixed effects models for our follow-up analyses of glucose, as variance was not equal across groups (Figure 3C & D). We saw no significant differences in glucose levels between people assigned male or female at birth in either OCC or PFC (*Χ*^2^_1_ values < 0.08, *p* values > 0.7). There were also no significant correlations between glucose levels in these regions and age (Spearman’s *r* values < 0.15, *p* values > 0.11).

We did observe a significant correlation between higher glucose levels in OCC and higher BMI (*r*_120_ = 0.24, *p* = 0.007), but this relationship was not significant in PFC (*r*_92_ = 0.14, *p* = 0.18). Correlations between glucose levels in OCC and BMI were comparable in magnitude (but not significant) within our group of relatives when considered alone (*r*_31_ = 0.28, *p* = 0.11), but were numerically lower in PwPP (*r*_50_ = 0.10, *p* = 0.5). This is notable as BMI, but not glucose levels in OCC, were significantly higher among relatives vs. controls (see main text).

Although we cannot rule out an influence of higher BMI on increased glucose levels in PwPP and / or relatives, BMI alone may not fully account for the observed group differences in brain glucose levels.

#### Tissue composition

We also examined correlations between metabolite levels and the proportion of GM, WM, and CSF within each individual’s VOI, to examine whether the differences in metabolites we observed might be related to differences in tissue composition (e.g., reduced GM in PwPP) [98,99]. In OCC, we found that the correlation between higher NAA and a higher normalized proportion of GM (Supplemental Equation 1) was significant (*r*_137_ = 0.38, *p* = 4 x 10^-6^). However, groups did not differ significantly in normalized GM (*X*^2^_2, n = 141_ = 4.70, *p* = 0.095), suggesting differences in brain structure between groups may not be sufficient to fully account for our observation of lower occipital NAA in relatives and PwPP. Further, NAA levels in OCC were not significantly correlated with the proportion of CSF within the VOI (*r*_137_ = 0.16, *p* = 0.057), and CSF proportion did not differ across groups (*X*^2^_2, n = 141_ = 0.08, *p* = 1.0).

We also found that higher NAAG levels in PFC were correlated with a lower proportion of GM (*r*_108_ = −0.26, *p* = 0.005), but not with CSF levels, *r*_108_ = 0.14, *p* = 0.14. However, once again we found no significant group differences between the proportions of GM (normalized) or CSF in PFC (*X*^2^_2_ values < 4.08, *p* values > 0.13). This suggests once more that group differences in brain structure may not be sufficient to account for higher NAAG levels in PFC in PwPP.

Higher glucose levels were correlated with higher levels of CSF within the VOI in both OCC (*r*_139_ = 0.19, *p* = 0.03) and PFC (*r*_110_ = 0.41, *p* = 9 x 10^-6^), as expected [69–71]. However, as noted above, we found no significant group differences in CSF fraction in either VOI (*Χ*^2^_2_ values < 3.2, *p* values > 0.2), suggesting that the amount of CSF within the VOI may not be sufficient to fully explain the group differences in glucose levels we observed (Figure 3C & D). We also saw no correlations between glucose levels and the normalized proportion of GM within the VOI (*r* values < 0.12, *p* values > 0.2).

#### Antipsychotic medication

Finally, we asked whether differences in NAA, NAAG, or glucose in PwPP may have been driven by antipsychotic medication. However, in PwPP, we found no significant associations between chlorpromazine equivalent doses [54,55] and NAA in OCC (*r*_56_ = −0.14, *p* = 0.3) or NAAG in PFC (*r*_48_ = −0.01, *p* = 1.0). Nor were there any significant correlations between antipsychotic medication scores and glucose levels in PwPP (*r* values < 0.05, *p* values > 0.7). Thus, we did not find strong evidence to suggest a direct link between antipsychotic medication levels and the observed group differences in brain metabolites.

#### VOI overlap across sessions

For participants who completed retest scans (10 controls, 36 PwPP), we calculated the overlap between VOIs across sessions using Dice coefficients. We found that VOI placement tended to overlap highly across sessions for both OCC (mean Dice coefficient = 0.78, *SD* = 0.15) and PFC (mean = 0.77, *SD* = 0.16).

---

**Supplemental Figure 1.**
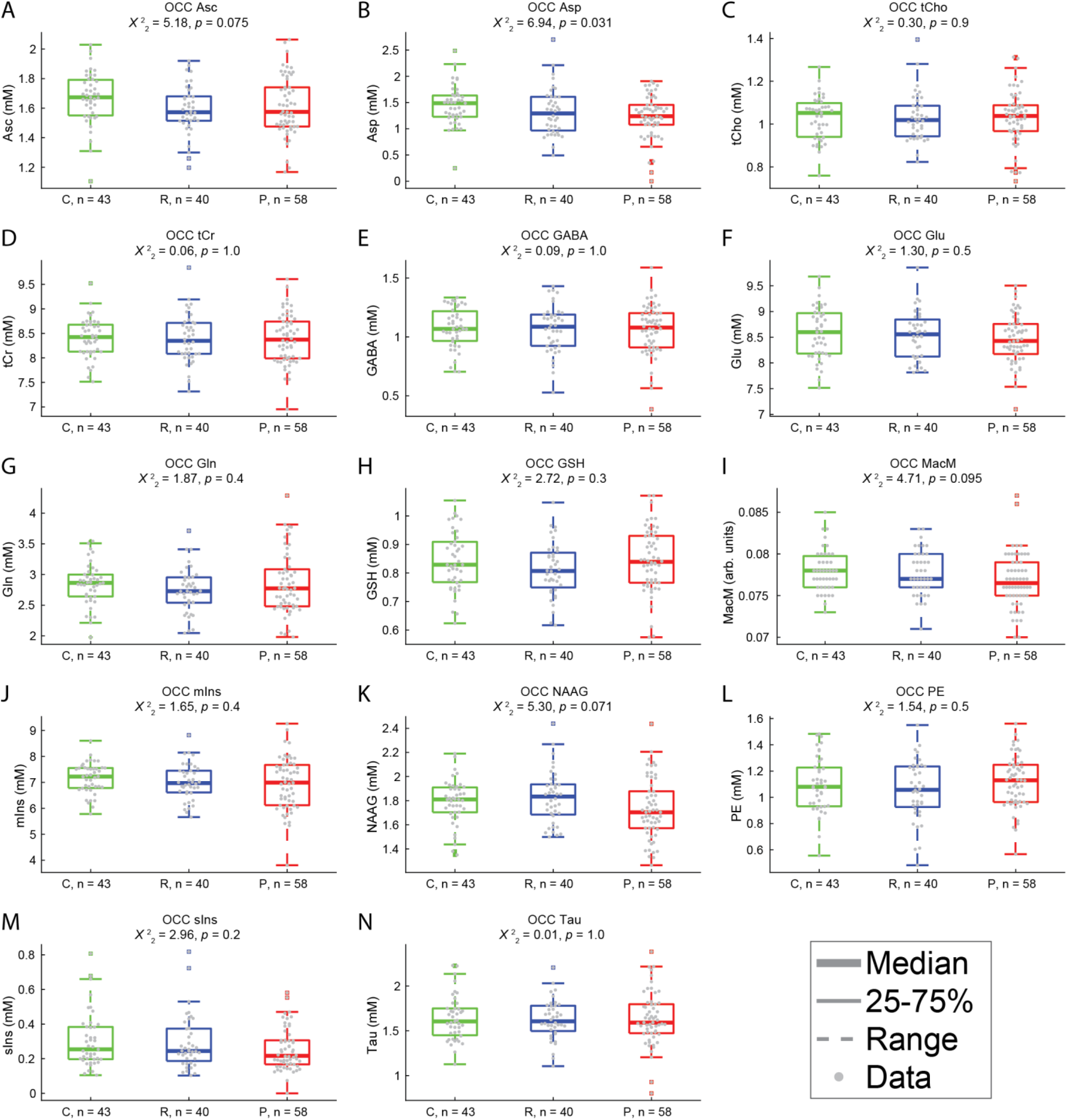
Additional metabolites in OCC. Uncorrected *p value*s are reported; group differences in these metabolites were either not significant (*a priori*; A-E) or did not survive correction for multiple comparisons (*post hoc*; F-N). C = controls, R = relatives, P = people with psychotic psychopathology. Thick lines show group medians, boxes show 25-75%, whiskers show 1.5x interquartile range, dots show individual data points. Repeat scan data are not shown. Metabolite abbreviations not defined in the main text are as follows: glutamate (Glu), glutamine (Gln), glutathione (GSH), ascorbic acid (Asc), aspartic acid (Asp), total choline (tCho), total creatine (tCr), glucose (Glc), macromolecules (MacM), *myo*-inositol (mIns), phosphorylethanolamine (PE), *scyllo*-inositol (sIns), taurine (Tau).

**Supplemental Figure 2.**
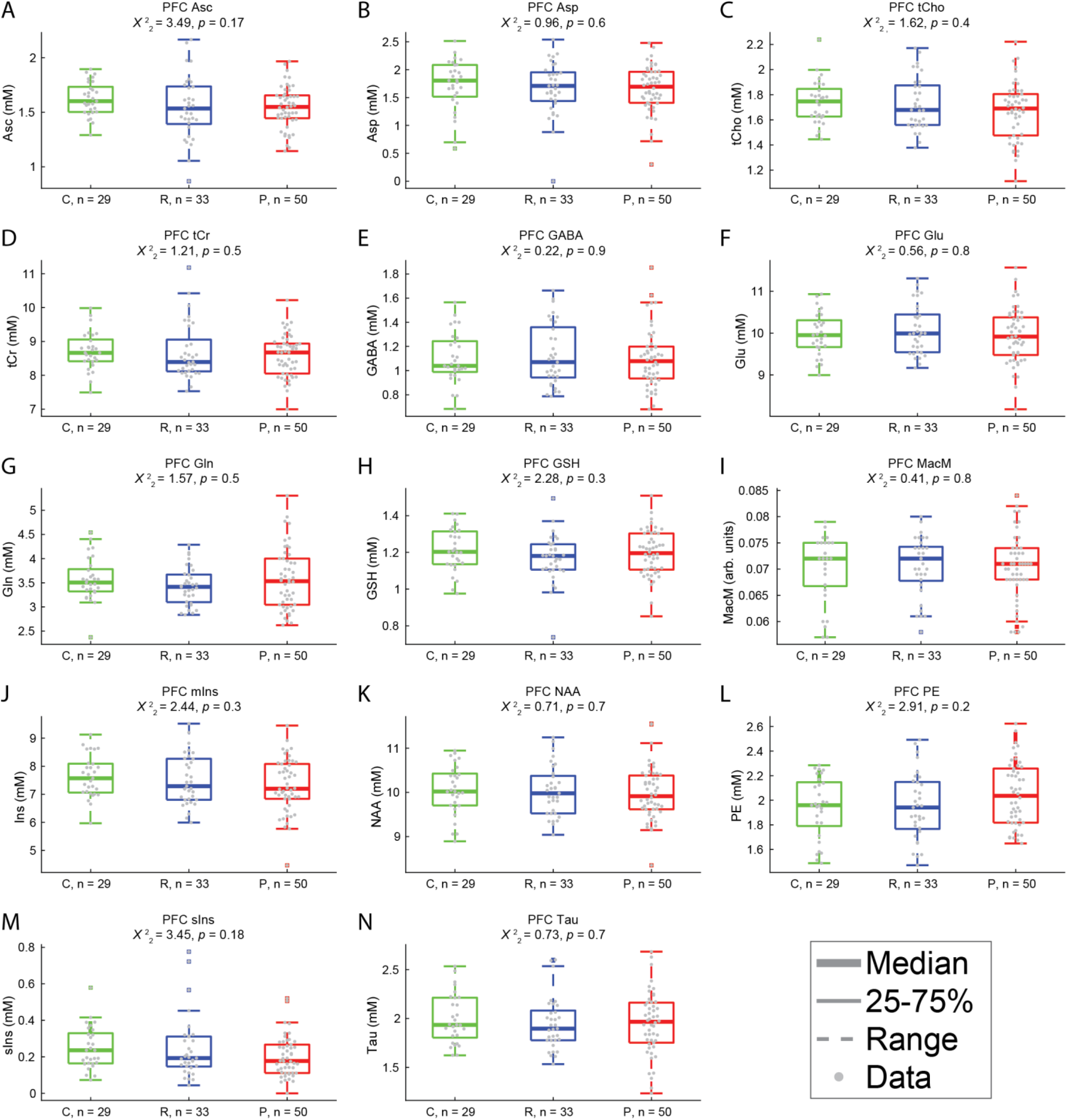
Additional metabolites in PFC. Uncorrected *p value*s are reported; group differences in these metabolites were either not significant (*a priori*; A-E) or did not survive correction for multiple comparisons (*post hoc*; F-N). C = controls, R = relatives, P = people with psychotic psychopathology. Thick lines show group medians, boxes show 25-75%, whiskers show 1.5x interquartile range, dots show individual data points. Repeat scan data are not shown. Metabolite abbreviations not defined in the main text are as follows: glutamate (Glu), glutamine (Gln), glutathione (GSH), ascorbic acid (Asc), aspartic acid (Asp), total choline (tCho), total creatine (tCr), glucose (Glc), macromolecules (MacM), *myo*-inositol (mIns), phosphorylethanolamine (PE), *scyllo*-inositol (sIns), taurine (Tau).

**Supplemental Figure 3.**
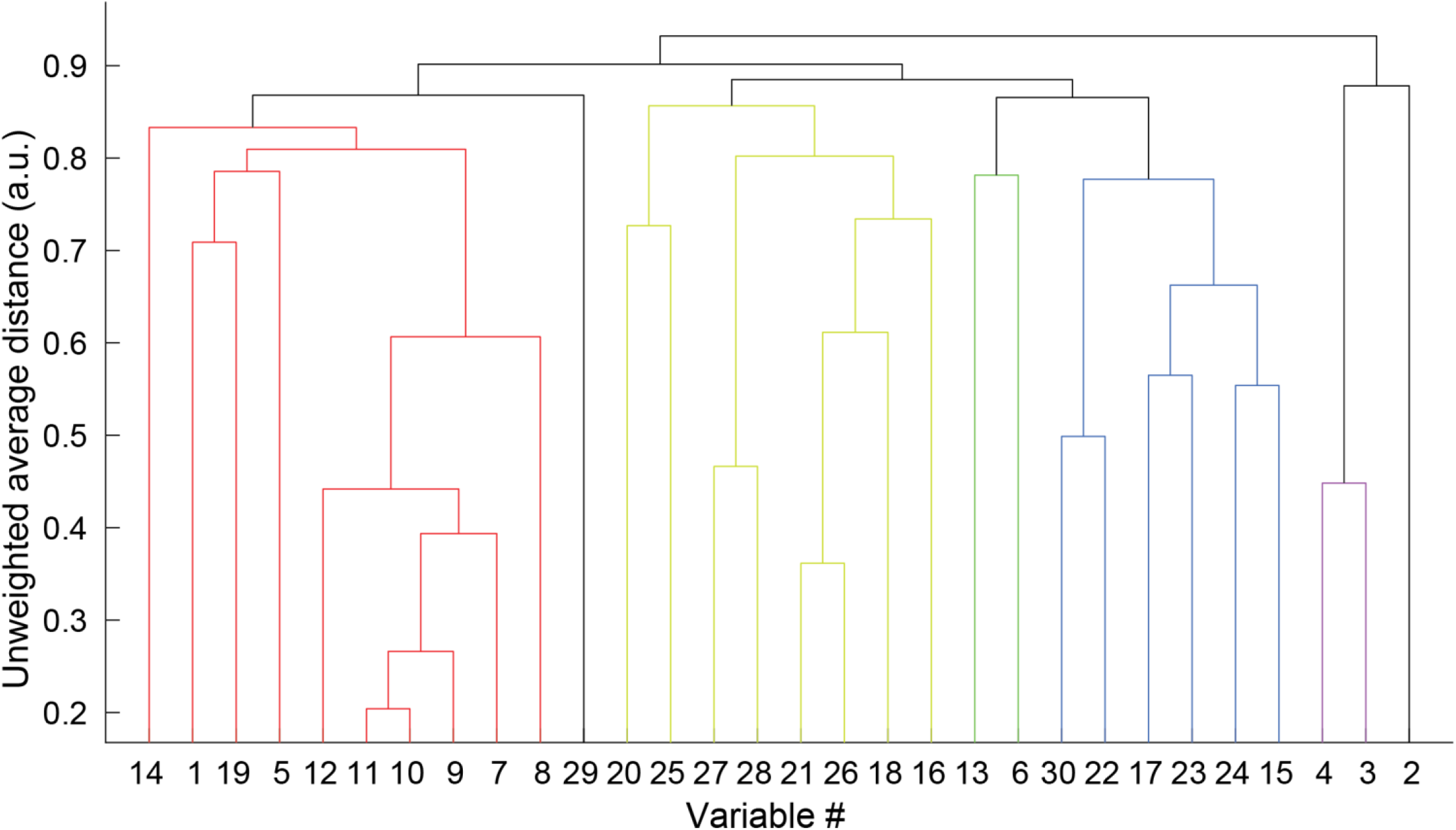
OCC hierarchical clustering dendrogram. The height of each bar indicates the distance between the clustered variables joined by that link. Colors indicate which of the 7 clusters each variable was included in (black indicates singleton clusters; see also Figure 5). Variable names are listed in Table 2.

**Supplemental Figure 4.**
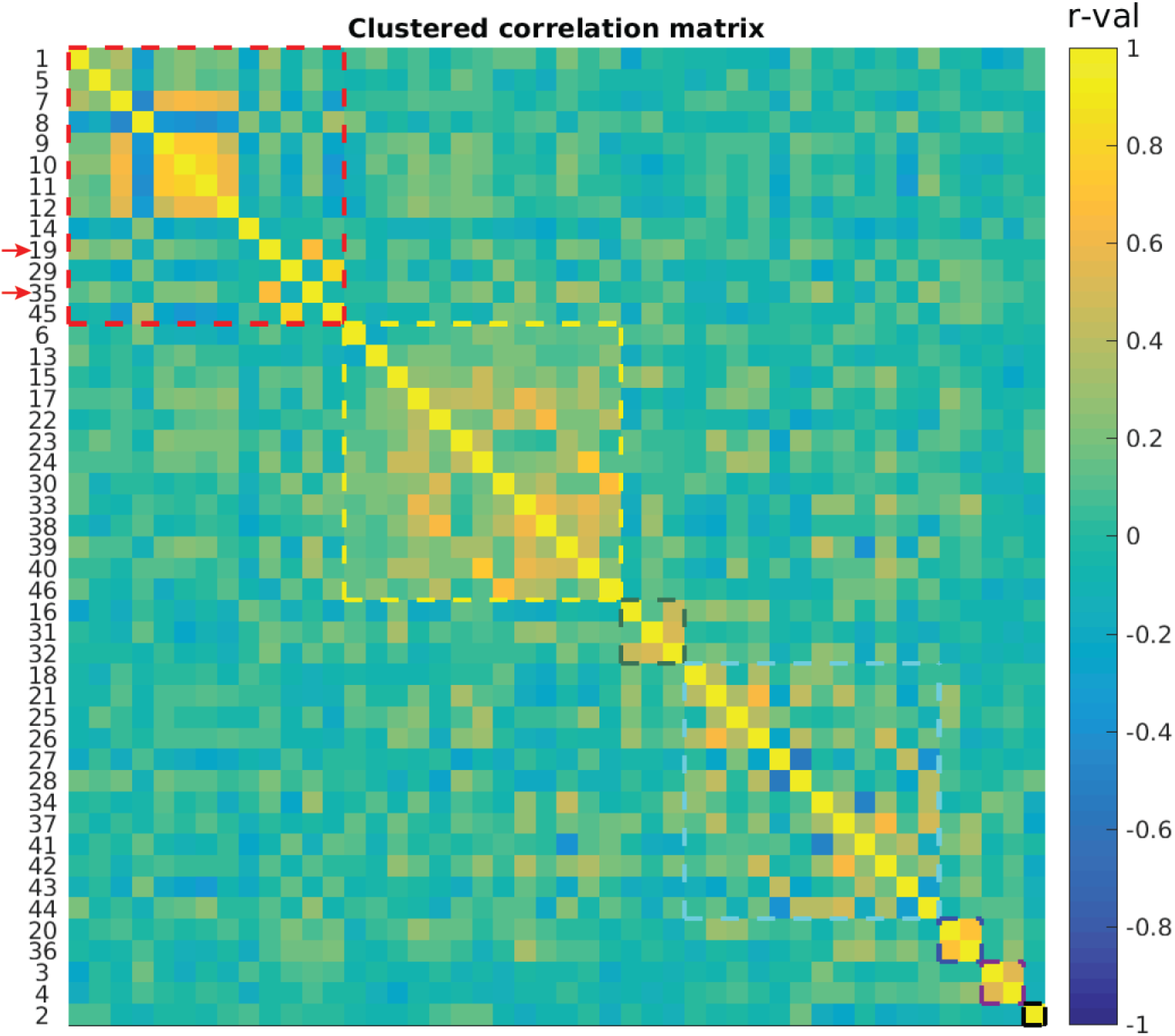
OCC+PFC hierarchical clustering results. Cells show pairwise Spearman correlation *r* values for measures from visual tasks (variables #1-7), clinical assessments (#8-14), MRS in OCC (#15-30) and PFC (#31-46). See Supplemental Table 2 for a full list of variables by number. Colored dashed lines indicate the boundaries of various clusters (*k* = 7). Red arrow indicates glucose in OCC (#19) and PFC (#35), which tended to cluster with visual task and clinical measures (red box).

**Supplemental Figure 5.**
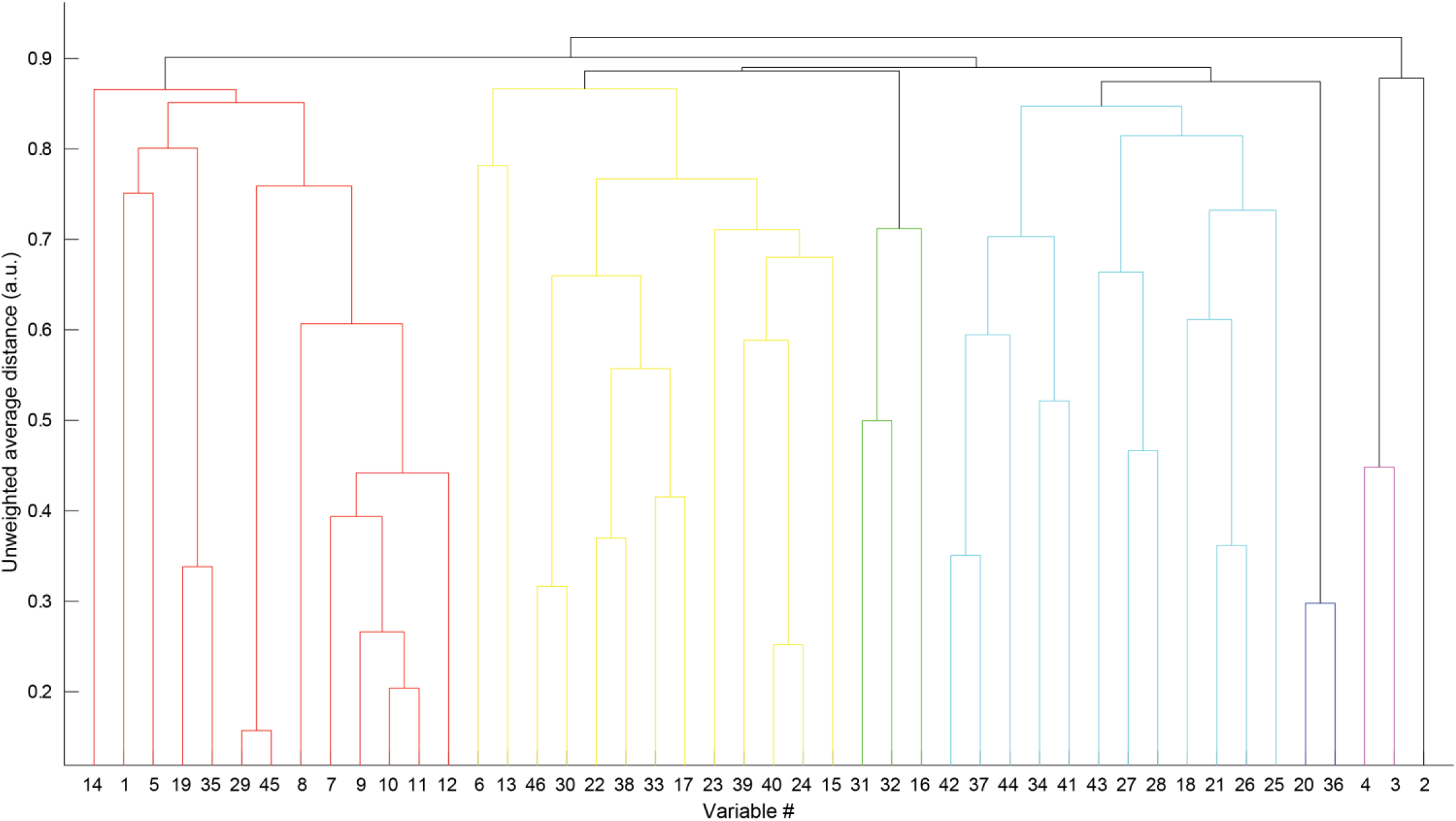
OCC+PFC clustering dendrogram. The height of each bar indicates the distance between the clustered variables joined by that link. Colors indicate which of the 7 clusters each variable was included in (black indicates singleton clusters; see also Supplemental Figure 4). Variable names are listed in Supplemental Table 2.

**Supplemental Table 1.** Water scaling correction parameters. Columns show the water content (%), *T*1 and *T*2 relaxation time constants (ms) for gray matter (GM), white matter (WM), and cerebrospinal fluid (CSF; rows). These values are based on previous work [61].

| | Water content | $T_1$ relaxation time constant (ms) | $T_2$ relaxation time constant (ms) |
| --- | --- | --- | --- |
| GM | 80% | 2130 | 50 |
| WM | 71% | 1220 | 55 |
| CSF | 97% | 4425 | 141 |

**Supplemental Table 2.** List of variables for OCC+PFC hierarchical clustering analysis.

| Variable # | Variable type | Variable name |
| --- | --- | --- |
| 1 | Visual task | Contrast discrimination threshold |
| 2 | Visual task | Contrast surround suppression index |
| 3 | Visual task | Contour object discrimination accuracy |
| 4 | Visual task | Contour object jitter threshold |
| 5 | Visual task | Structure-from-motion bi-stable switch rate |
| 6 | Visual task | Structure-from-motion bi-stable coefficient of variance |
| 7 | Clinical measure | BPRS total score |
| 8 | Clinical measure | BACS Z-score |
| 9 | Clinical measure | SGI total score |
| 10 | Clinical measure | SPQ total score |
| 11 | Clinical measure | PID-5 psychoticism factor |
| 12 | Clinical measure | BPRS hallucination score |
| 13 | Clinical measure | Snellen visual acuity (decimal fraction) |
| 14 | Clinical measure | Mars visual contrast sensitivity |
| 15 | MRS measure | OCC ascorbic acid |
| 16 | MRS measure | OCC aspartic acid |
| 17 | MRS measure | OCC total creatine |
| 18 | MRS measure | OCC GABA |
| 19 | MRS measure | OCC glucose |
| 20 | MRS measure | OCC glutamine |
| 21 | MRS measure | OCC glutamate |
| 22 | MRS measure | OCC total choline |
| 23 | MRS measure | OCC glutathione |
| 24 | MRS measure | OCC *myo*-inositol |
| 25 | MRS measure | OCC macromolecules |
| 26 | MRS measure | OCC NAA |
| 27 | MRS measure | OCC NAAG |
| 28 | MRS measure | OCC phosphorylethanolamine |
| 29 | MRS measure | OCC *scyllo*-inositol |
| 30 | MRS measure | OCC taurine |
| 31 | MRS measure | PFC ascorbic acid |
| 32 | MRS measure | PFC aspartic acid |
| 33 | MRS measure | PFC total creatine |
| 34 | MRS measure | PFC GABA |
| 35 | MRS measure | PFC glucose |
| 36 | MRS measure | PFC glutamine |
| 37 | MRS measure | PFC glutamate |
| 38 | MRS measure | PFC total choline |
| 39 | MRS measure | PFC glutathione |
| 40 | MRS measure | PFC *myo*-inositol |
| 41 | MRS measure | PFC macromolecules |
| 42 | MRS measure | PFC NAA |
| 43 | MRS measure | PFC NAAG |
| 44 | MRS measure | PFC phosphorylethanolamine |
| 45 | MRS measure | PFC *scyllo*-inositol |
| 46 | MRS measure | PFC taurine |

**Supplemental Table 3.** Metabolite concentrations. Data are reported as means and standard deviations (SD) for occipital (OCC) and prefrontal (PFC) VOIs in controls (n = 43 in OCC, n = 29 in PFC), relatives (n = 40 in OCC, n = 33 in PFC), and PwPP (n = 58 in OCC, n = 50 in PFC). Units are millimolar (mM), except for macromolecules (arbitrary units).

| Metabolite | Controls,<br>mean | Controls,<br>SD | Relatives,<br>mean | Relatives,<br>SD | PwPP,<br>mean | PwPP,<br>SD |
| --- | --- | --- | --- | --- | --- | --- |
| OCC ascorbic acid | 1.68 | 0.18 | 1.59 | 0.16 | 1.61 | 0.20 |
| OCC aspartic acid | 1.40 | 0.38 | 1.32 | 0.45 | 1.20 | 0.40 |
| OCC total choline | 1.03 | 0.10 | 1.03 | 0.11 | 1.04 | 0.13 |
| OCC total creatine | 8.43 | 0.40 | 8.40 | 0.50 | 8.38 | 0.51 |
| OCC GABA | 1.06 | 0.17 | 1.07 | 0.20 | 1.07 | 0.23 |
| OCC glucose | 1.13 | 0.43 | 1.19 | 0.65 | 1.38 | 0.57 |
| OCC glutamate | 8.56 | 0.50 | 8.54 | 0.47 | 8.47 | 0.45 |
| OCC glutamine | 2.84 | 0.36 | 2.74 | 0.36 | 2.86 | 0.53 |
| OCC glutathione | 0.85 | 0.10 | 0.81 | 0.09 | 0.84 | 0.11 |
| OCC macromolecules | 0.078 | 0.003 | 0.078 | 0.003 | 0.077 | 0.004 |
| OCC *myo*-inositol | 7.15 | 0.58 | 7.01 | 0.69 | 6.92 | 0.98 |
| OCC NAA | 11.00 | 0.57 | 10.81 | 0.65 | 10.81 | 0.49 |
| OCC NAAG | 1.80 | 0.18 | 1.83 | 0.22 | 1.75 | 0.23 |
| OCC phosphorylethanolamine | 1.10 | 0.20 | 1.05 | 0.23 | 1.10 | 0.21 |
| OCC *scyllo*-inositol | 0.30 | 0.15 | 0.29 | 0.16 | 0.25 | 0.12 |
| OCC taurine | 1.63 | 0.26 | 1.62 | 0.22 | 1.65 | 0.28 |
| PFC ascorbic acid | 1.62 | 0.15 | 1.54 | 0.29 | 1.55 | 0.20 |
| PFC aspartic acid | 1.73 | 0.46 | 1.67 | 0.49 | 1.69 | 0.42 |
| PFC total choline | 1.74 | 0.17 | 1.73 | 0.21 | 1.68 | 0.22 |
| PFC total creatine | 8.72 | 0.56 | 8.68 | 0.83 | 8.52 | 0.61 |
| PFC glucose | 1.03 | 0.51 | 1.17 | 0.65 | 1.46 | 0.69 |
| PFC GABA | 1.10 | 0.21 | 1.13 | 0.26 | 1.08 | 0.24 |
| PFC glutamate | 9.98 | 0.51 | 10.07 | 0.59 | 9.89 | 0.65 |
| PFC glutamine | 3.57 | 0.45 | 3.43 | 0.39 | 3.56 | 0.63 |
| PFC glutathione | 1.21 | 0.12 | 1.17 | 0.13 | 1.18 | 0.12 |
| PFC macromolecules | 0.070 | 0.006 | 0.071 | 0.006 | 0.071 | 0.006 |
| PFC *myo*-inositol | 7.64 | 0.75 | 7.50 | 0.90 | 7.31 | 0.94 |
| PFC NAA | 10.05 | 0.56 | 10.03 | 0.57 | 9.88 | 0.65 |
| PFC NAAG | 1.26 | 0.18 | 1.29 | 0.19 | 1.18 | 0.18 |
| PFC phosphorylethanolamine | 1.94 | 0.25 | 1.95 | 0.27 | 2.02 | 0.26 |
| PFC *scyllo*-inositol | 0.25 | 0.12 | 0.25 | 0.17 | 0.20 | 0.11 |
| PFC taurine | 2.00 | 0.25 | 1.95 | 0.27 | 1.93 | 0.29 |

**Supplemental Table 4.** Longitudinal variability in metabolite concentrations. Intra-class correlation coefficients (ICC(3,k) [67]) were used to quantify variability in metabolite concentrations across two longitudinal scanning sessions collected in a subset of participants (10 controls, 36 PwPP, see Table 1 for information about the amount of time between scanning sessions). Note that of the 36 PwPP who completed repeat scanning sessions, OCC data from one or both sessions were excluded for 4 individuals. Likewise, PFC data were excluded for 2 PwPP who completed repeat sessions.

| **Metabolite** | **ICC in OCC**<br>n = 10 controls,<br>32 PwPP | **ICC in PFC**<br>n = 26 PwPP |
| --- | --- | --- |
| ascorbic acid | 0.83 | 0.47 |
| aspartic acid | 0.77 | 0.64 |
| total choline | 0.86 | 0.91 |
| total creatine | 0.89 | 0.84 |
| GABA | 0.62 | 0.63 |
| glucose | 0.56 | 0.67 |
| glutamate | 0.79 | 0.80 |
| glutamine | 0.92 | 0.92 |
| glutathione | 0.78 | 0.44 |
| macromolecules | 0.69 | 0.62 |
| *myo*-inositol | 0.95 | 0.95 |
| NAA | 0.83 | 0.74 |
| NAAG | 0.82 | 0.78 |
| phosphorylethanolamine | 0.75 | 0.85 |
| *scyllo*-inositol | 0.96 | 0.89 |
| taurine | 0.89 | 0.76 |

## Notes

### Competing Interest Statement

The authors have declared no competing interest.

### Author Declarations

IRB of The University of Minnesota gave ethical approval for this work

